# Contextual Adaptation and Implementation of WHO Guideline on Self-Care Interventions for SRH in Kenya, Nigeria and Uganda

**DOI:** 10.1101/2025.05.06.25327056

**Authors:** Austen El-Osta, Eva Riboli Sasco, Aos Alaa, Manisha Karki, Andrea Cutherell, Amanda Kalamar, Ricky Banarsee, Azeem Majeed

## Abstract

**Background:** Self-care interventions for sexual and reproductive health and rights (SRHR) are critical to advancing individual wellbeing and achieving universal health coverage. This study assesses how three countries - Kenya, Nigeria and Uganda - have adapted and implemented the World Health Organization (WHO) Guideline on Self-Care Interventions for SRHR within their national policy and practice landscapes.

**Objectives:** The primary objective was to develop and pilot a novel policy mapping and implementation analysis tool. Secondary aims included using a mixed-methods approach comprising policy document review, surveys and interviews to evaluate the contextualisation and uptake of WHO recommendations at the country level.

**Methods:** We designed a Policy Mapping and Implementation Matrix (PMIM) to assess alignment with 24 WHO SRHR self-care recommendations. Data were collected from 316 stakeholders through surveys and interviews, and 47 policy documents were reviewed. Findings were synthesised using a Red-Amber-Green (RAG) matrix to assess implementation across the guideline’s five domains.

**Results:** Implementation varied by country and recommendation. Family planning and infertility services (Category 2) showed the strongest uptake, while areas such as unsafe abortion management and STI self-sampling (Categories 3 and 4) were less consistently addressed. Kenya demonstrated broad alignment through multiple policies, while Nigeria and Uganda showed promising progress, particularly with the development of dedicated national self-care guidelines. Key barriers included supply chain challenges, low health literacy and legal constraints; critical enablers were provider training and task shifting.

**Conclusion:** This study introduces a novel, pragmatic framework for assessing national self-care policy and practice. It highlights the importance of contextual adaptation rather than mechanical adoption of global guidelines. While study limitations are acknowledged, the methodology offers a replicable approach for monitoring and strengthening self-care integration in diverse settings.

**Key points:**

- Little is known about how countries are contextualising and adapting WHO Guideline on Self-Care interventions
- We present the first iteration policy mapping and implementation analysis tool and findings from three countries where there is an emergent self-care movement
- A key driver for contextual adaptation of the WHO Guideline includes the development of national guidelines and self-care advocacy groups
- The extent that the policy landscape supported self-care practices for SRHR could be used to objectively prioritise policy and advocacy opportunities in each country

## Introduction

Self-care has emerged as a transformative paradigm in global health, particularly within the domain of sexual and reproductive health and rights (SRHR). By enhancing individual agency and autonomy, self-care interventions offer new pathways for people to manage their health, especially in contexts marked by systemic inequities and constrained health systems (1, 2). Recognising this, the World Health Organization (WHO) published the Consolidated Guideline on Self-Care Interventions for SRHR in 2019 (3). This landmark guidance outlines 24 evidence-based recommendations across five categories, ranging from antenatal and postnatal care to family planning, prevention of unsafe abortion, management of sexually transmitted infections (STIs) and promotion of reproductive rights (**Figure 1**).

**Figure 1:**
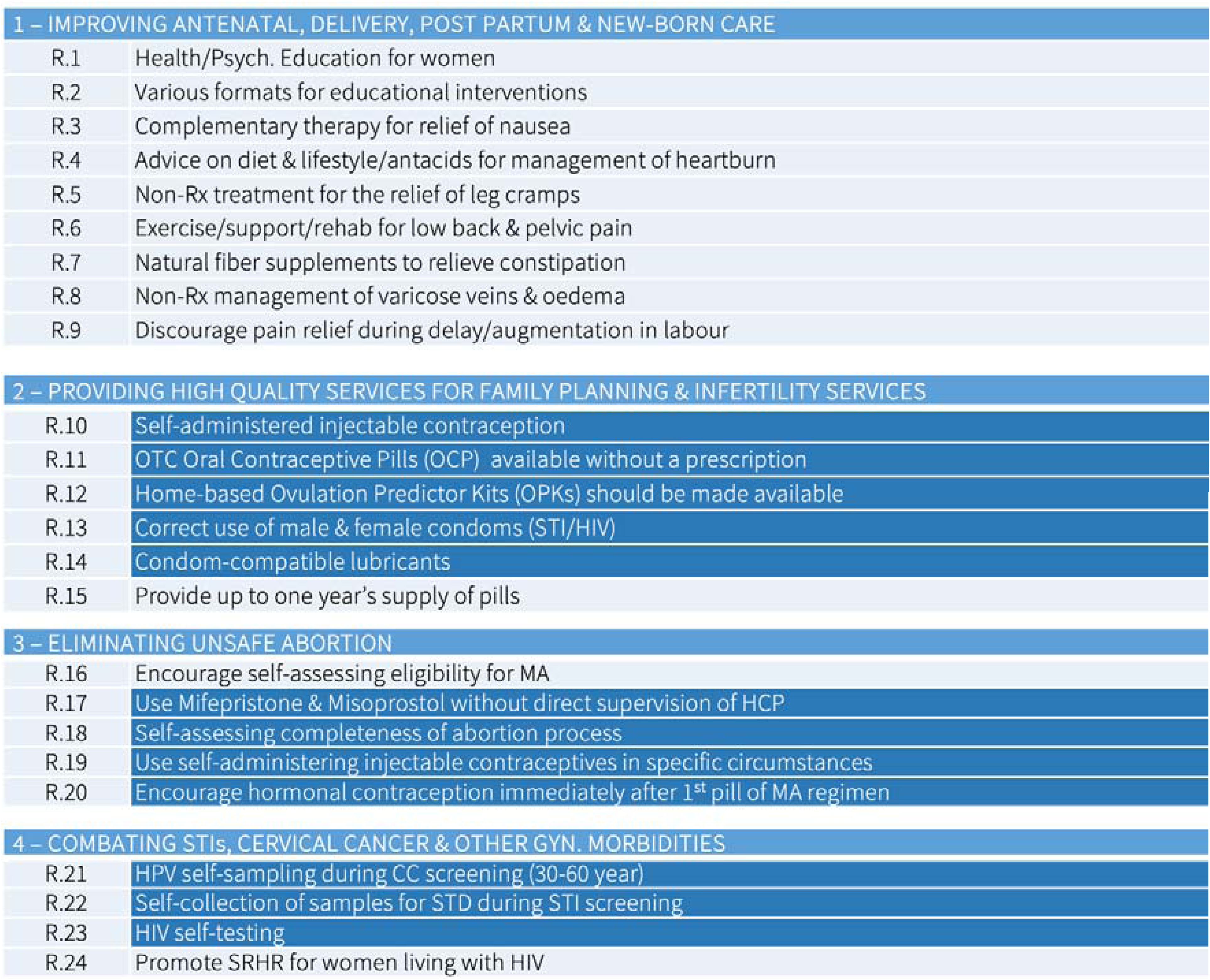
Overview of WHO self-care recommendations for sexual and reproductive health (abridged)

Self-care transcends conventional healthcare boundaries (4, 5), providing a transformative lens through which to position, utilise and operationalize SRHR products and services. The guideline highlights that the successful uptake of self-care interventions hinges on enabling environments both at policy and system levels that support individual decision-making and access to safe, effective tools and services. It includes both clinical and operational recommendations, along with programmatic guidance to inform country-level adaptation. Crucially, it positions self-care not as a substitute for formal health systems, but as a complementary approach that can extend the reach of services, especially for underserved populations.

Since the guideline’s publication, and subsequent update in 2022 (6) there has been growing interest in how countries are contextualising and integrating its recommendations. Yet to date, no validated framework exists to assess national-level alignment with the WHO guidance. Moreover, little is known about how implementation varies by context, or the factors that facilitate or hinder adoption in different health policy environments.

To address this gap, we undertook a mixed-methods study to examine how three countries (Kenya, Nigeria and Uganda) are adapting and implementing the WHO self-care recommendations for SRHR. These countries were selected due to their early engagement with self-care movements and emerging national policy frameworks. The primary aim of this study was to develop and pilot a pragmatic policy mapping and implementation tool. By synthesising data from national policy documents, stakeholder surveys and key informant interviews, we sought to provide a comprehensive and comparative analysis of self-care policy and practice in these distinct settings.

## Methods

This mixed-methods study was designed to evaluate how national policies in Kenya, Nigeria and Uganda align with the WHO *Guideline on Self-Care Interventions for SRHR*. The study comprised fouu main components: (i) policy mapping, (ii) stakeholder mapping, (iii) data collection via electronic survey and interviews, and (iv) development and application of a novel policy mapping and implementation matrix (PMIM). Our methodological approach was informed by established frameworks in policy research and social inquiry, including the *Rapid Policy Network Mapping* (RPNM) methodology developed by Bainbridge et al., which offers a structured means of identifying and analysing governance actors, networks and policy instruments within a defined policy space (7, 8).

### Policy mapping

We adopted a rapid scoping methodology to assess national policy environments related to self-care for SRHR. Academic literature, grey literature and national policy documents and White Papers from Kenya, Nigeria and Uganda were identified via desktop research and consultations with in-country stakeholders. Inclusion was limited to English-language documents from government or quasi-governmental sources. Policy documents were reviewed by a team of five researchers to extract “control statements” - text segments reflecting alignment with the WHO’s 24 self-care recommendations. Relevant documents published between 2010-2019 were included.

A bespoke policy mapping and implementation matrix (PMIM) was developed to systematically document two key aspects: (i) evidence highlighting the level of contextualization, adaptation, or implementation for each recommendation, and (ii) the corresponding policy reference linked to the appropriate cell for each of the 24 recommendations within each country.

PMIM provided a structured format for documenting the presence or absence of relevant control statements, mapped to each of the 24 WHO recommendations across the three countries. A Red-Amber-Green (RAG) traffic light coding scheme was employed to categorise implementation intensity where Green signalled high levels of adaptation or implementation, Amber signalled moderate levels and Red signalled low or no alignment. Cells were left blank where no relevant policy content was found.

### Stakeholder mapping

To comprehensively assess the landscape of self-care for SRHR, a stakeholder mapping approach was employed. This approach aimed to identify and categorize the key stakeholders involved in self-care policy development, implementation and advocacy within the selected countries of Kenya, Nigeria and Uganda. Stakeholder identification was based on a multi-step process that included desk research, consultation with local experts and engagement with relevant organizations. Initially, an extensive review of policy documents, white papers and academic literature was conducted to identify potential stakeholders. This was followed by consultation with in-country experts, including healthcare professionals, researchers and representatives from civil society organizations, to validate and expand the list of stakeholders.

The identified stakeholders were then categorized into different groups based on their roles, interests and influence in the field of SRHR self-care. These categories included government entities, non-governmental organizations (NGOs), healthcare providers, professional associations, advocacy groups, academia and international organizations. This stakeholder mapping process provided a comprehensive overview of the various actors engaged in shaping self-care policies and practices within each country.

Stakeholder engagement was further facilitated through surveys, interviews and focus group discussions, enabling deeper insights into their perspectives, experiences and contributions to self-care implementation. The stakeholder mapping approach served as a foundational step to understand the landscape of self-care for SRHR whereas engagement with diverse stakeholders enriched the analysis of policy and practice within the study countries.

### Electronic survey

We conducted a cross-sectional online survey of self-care stakeholders in each country. The link to the electronic survey was published and made available on the Imperial College London Qualtrics platform between 14 December 2020 and 5 March 2021 (12 weeks). The survey was open and could be accessed by anyone with a link. We also used semi-structured interviews to surface assumptions from a convenience sample of self-care stakeholders from each country. For the interviews, we selected a sub-sample to incorporate a mix of clinical roles and specialist interest in self-care practices for SRHR.

### Participant recruitment

Potentially eligible participants received an invitation email from the study team, including the participant information sheet (PIS) and a link to the survey. Eligible participants were 18 years or over, consented to take part in the research, agreed to complete the survey and/or interview and were English speaking.

The researchers’ personal and professional networks were mobilized to respond and further disseminate the eSurvey among eligible participants. The PIS included information regarding the study aims, the protection of participants’ personal data, their right to withdraw from the study at any time, which data were stored, where and for how long, who the principal investigator was and the survey length. Participants were informed that this was a voluntary survey and monetary incentives were not offered but participants were advised that they could access the findings at a later stage whilst underlying the potential collective benefits of taking part in terms of helping advance knowledge in this area and the formulation of future policy and advocacy opportunities to promote self-care praxis. The data collected were stored on a password protect database hosted on Imperial College London encrypted server that can only be accessed by the research team.

### Online tool

The modular survey comprised a total of 79 items displayed on one page and was accessible using a personal computer or smartphone. The full survey is included in **Supplementary file 1**. We used the online tool to collect semi-quantitative data using multiple choice and Likert scale from a wide mix of respondents to gauge awareness of operative policies, perception around the levels of availability of SRHR products and the extent that each recommendation in the WHO guideline was being implemented, contextualised or adapted in policy and practice. Questions regarding demographic characteristics of the users included information on gender, age, ethnicity, educational level, country and professional role. Participants could review their answers before submitting them. All data collected through the survey were anonymised and not personally identifiable. The online survey technical functionality was tested before being published. The first question asked participants to confirm their consent to participate and this was followed by a filter question to identify the country. Experiences and perceptions related to the current availability of SRHR products were evaluated through several questions. Product availability was reported as either widely available, somewhat available, somewhat unavailable, or largely unavailable. Respondents were able to refrain from providing an answer by selecting ‘no opinion’ or ‘unsure’. Such answers were treated as missing data in all the analyses (listwise exclusion). We gave respondents the opportunity to indicate which categories of recommendations they would like to provide feedback on, given their own knowledge and expertise, so that only relevant questions appeared in the main (modular) survey. Questions concerning respondent’s perceptions as to the extent that each of the WHO recommendations was being adapted, contextualised or implemented, were presented on a matrix where respondents could indicate the perceived level of implementation. We also included 8 questions (2 per category) that invited free-text responses to capture contextual feedback as regards the main drivers and barriers to further alignment with each category of recommendations.

### Qualitative interviews

At the end of the survey, respondents could volunteer for a follow-up interview by submitting contact details. Those selected were sent a Participant Information Sheet and consent form. Interviews were conducted via Zoom or Microsoft Teams and lasted between 35–62 minutes. After consent was sought, all interviews were recorded, transcribed verbatim and analysed using thematic framework approach. An open coding approach was initially applied to transcripts by a single coder (ERS), with regular discussions held with the principal investigator (AEO) to develop a shared coding framework. Emergent themes were iteratively refined and mapped against the WHO guideline categories.

## Data Analysis

Quantitative survey data were summarised using frequencies and percentages. Incomplete surveys (n=15) were excluded from analysis. Data were stratified by country. Free-text responses and interview transcripts were thematically analysed to identify recurring themes, contextual drivers and implementation barriers.

Findings from policy mapping, survey responses and interviews were triangulated to populate the PMIM and assign RAG scores for each recommendation per country. This mixed-methods triangulation ensured a robust assessment of alignment with WHO guidance.

### Tool Development: Policy Mapping and Implementation Matrix (PMIM)

PMIM was developed as a structured tool to capture the extent of national-level contextualisation of WHO recommendations. Each recommendation was assessed across three data sources (i) Policy analysis which considered the presence of control statements in national documents, (ii) Survey data that captured information on stakeholder perceptions of implementation, and (iii) Interview data with qualitative insights on barriers and facilitators. The combined data were used to assign RAG ratings. Green indicated strong evidence of policy adoption and implementation; amber indicated partial or uneven implementation; red indicated limited or no evidence of action. This matrix allowed for country-specific and cross-country comparisons. This mixed methods approach allowed for a detailed exploration of the extent to which each recommendation was addressed and integrated within the policies and frameworks of Kenya, Nigeria and Uganda. The Checklist for Reporting Results of Internet e-Surveys (CHERRIES) (9), the Consolidated Criteria for Reporting Qualitative Research (COREQ), and the Mixed Methods Article Reporting Standards (MMARS) checklists (10) were used to improve the quality of the reporting.

### Ethics

The study received a favourable opinion from imperial college research ethics committee (#20IC6438). Participants consented to take part in the survey and personal interview.

### Patient and public involvement

No patient was involved. The study protocol and online survey were developed in collaboration with the Self-Care Trailblazer Group and national self-care networks in Kenya, Nigeria and Uganda.

## Results

### Policy landscape overview

A total of 47 policy documents and white papers were identified and reviewed: 8 from Kenya, 11 from Nigeria and 28 from Uganda **(Table 1**). A brief description of each asset is included in **Supplementary Table 2).** Of these, nine were found to be particularly relevant to the implementation of self-care for SRHR and contained control statements that aligned with WHO recommendations (see **Table 2**).

**Table 1:**
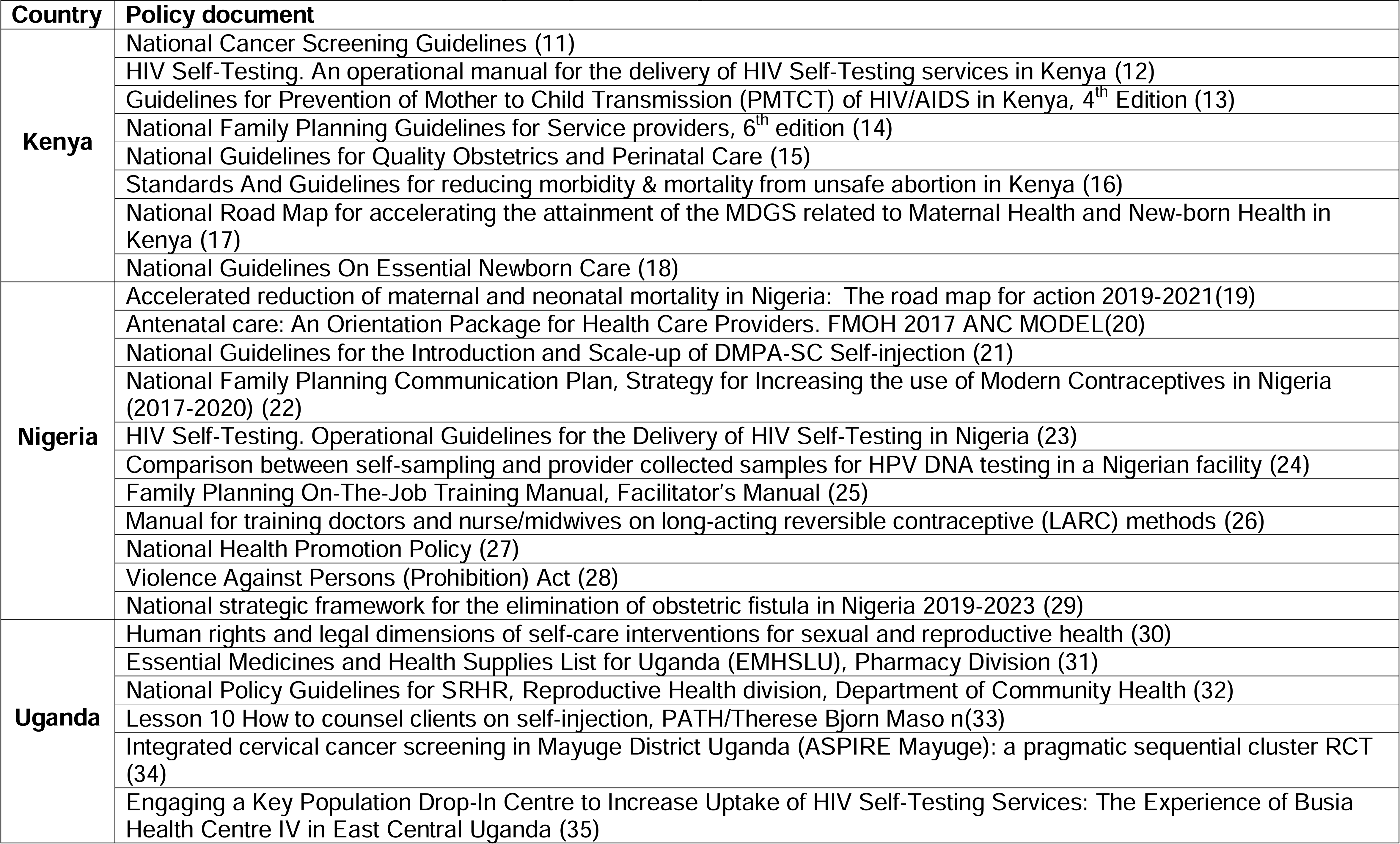
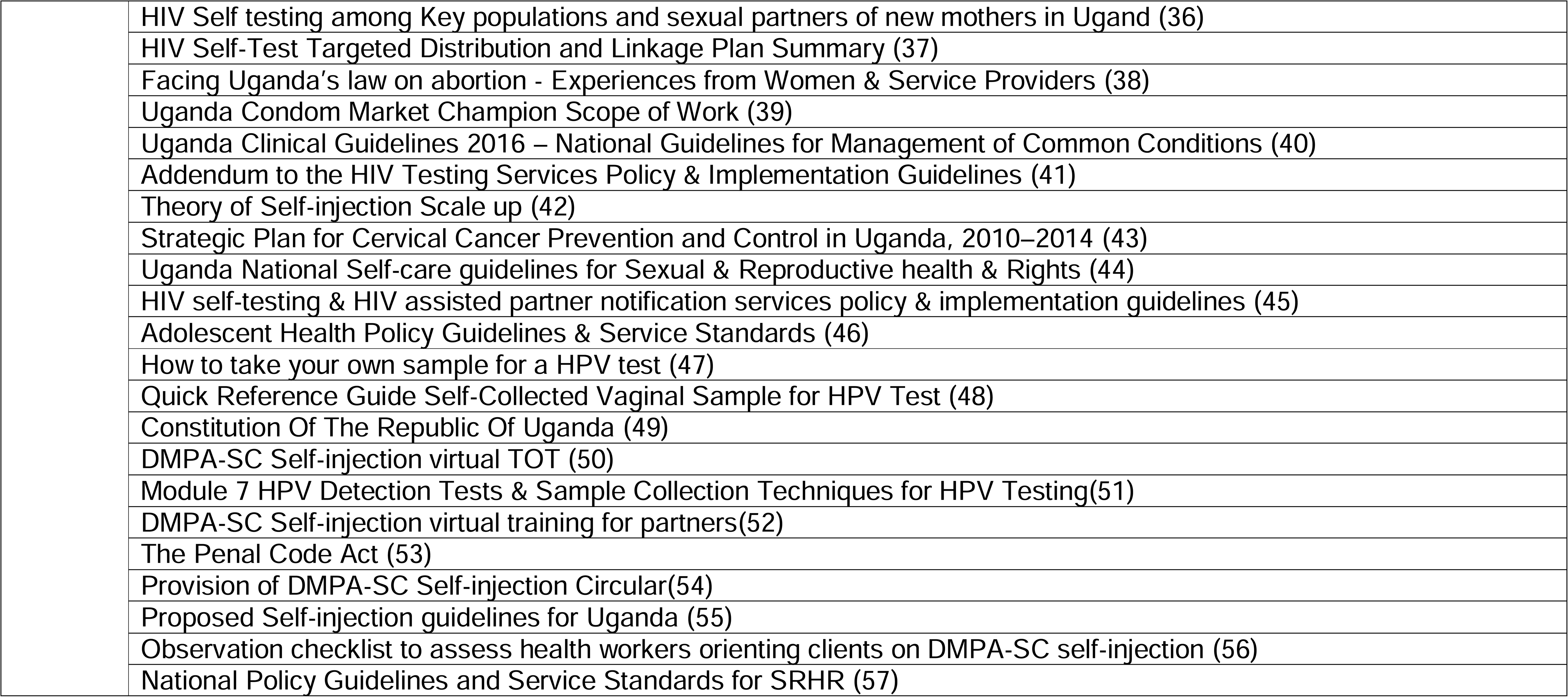
Table of assets identified from Kenya, Nigeria and Uganda.

**Table 2:**
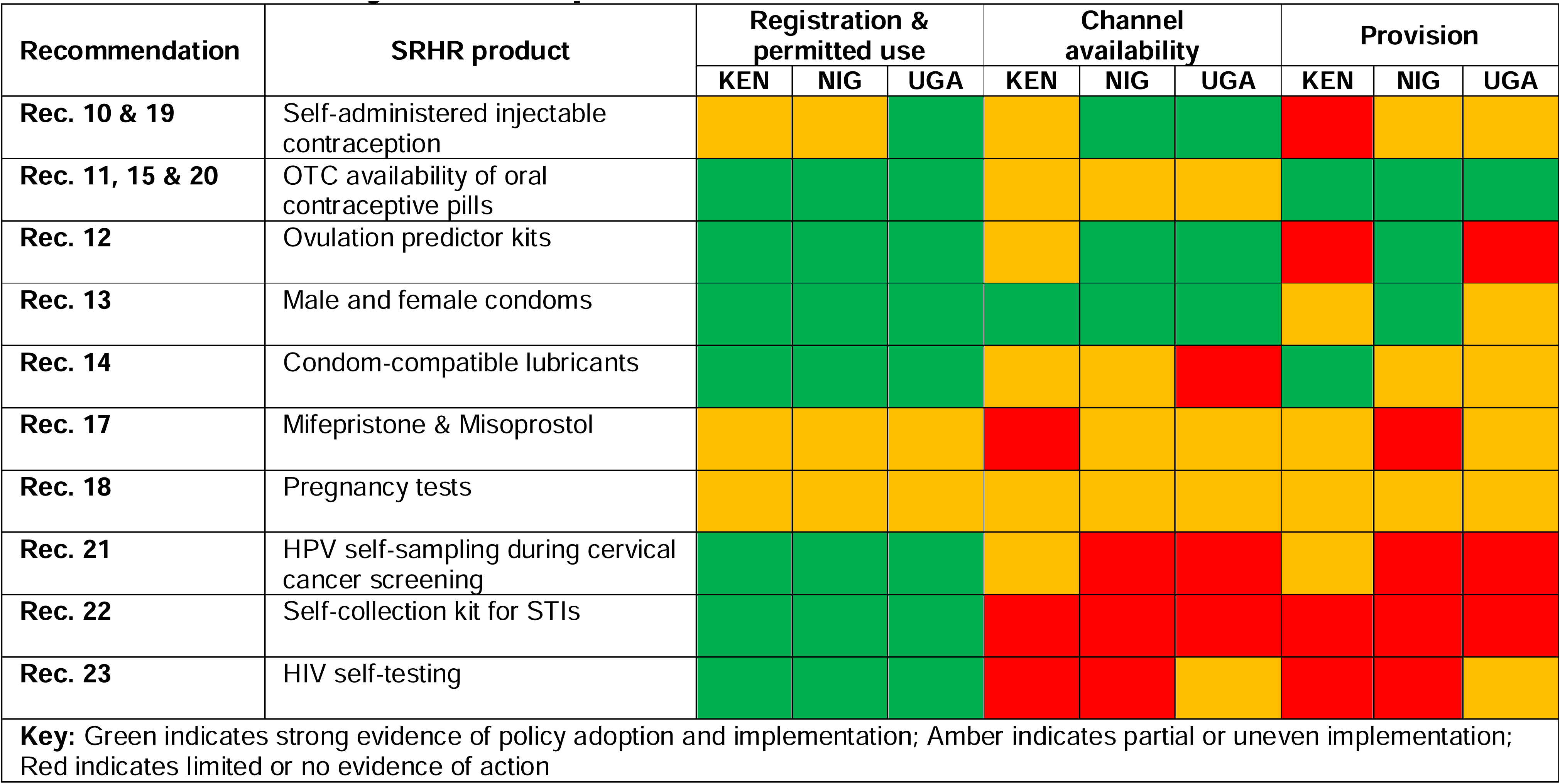
Availability of SRHR products relevant to recommendations.

Uganda’s National Self-Care Guidelines for Sexual and Reproductive Health and Rights (2020) and Nigeria’s National Guidelines on Self-Care for Sexual, Reproductive and Maternal Health (2021) were especially notable as they incorporated 92% (22/24) and 83% (20/24) of the WHO recommendations, respectively. Kenya, in the absence of a national self-care guideline, demonstrated alignment through control statements embedded in multiple existing documents, published between 2010 and 2018.

### Respondent characteristics

Only 76 /211 stakeholders invited to participate responded to the online survey. More than half (n=35) worked in NGOs, 15 were public health professionals, 5 worked in sexual health clinics, 2 worked in community pharmacy setting, 5 worked in academia, 2 were policy makers, and 25 did not disclose their designation or field of work. Forty percent of respondents completed information about the Kenya context, 40 from Uganda and 20 from Nigeria).

For the qualitative arm of the study, a total of 10 key informant interviews were conducted - three each from Kenya and Nigeria and four from Uganda. Interviewees provided nuanced insights into barriers and drivers for policy adaptation and implementation. The consolidated findings from respondent feedback are presented below.

### SRHR Product Availability

Survey findings (n=56) highlighted important variations in the perceived availability and delivery of SRHR commodities. Emergency contraception, male condoms and ovulation predictor kits were considered widely available across all three countries. However, access to DMPA-SC (self-injectable contraception), female condoms and lubricants was inconsistent. Uganda had the highest reported availability of DMPA-SC (78%), followed by Nigeria (50%) and Kenya (33%). Self-sampling kits for HPV and STIs were reported as least available, with only 17%-38% of respondents confirming access. HIV self-testing kits were more widely known and integrated into policies and campaigns, especially in Uganda. **Table 2** summarises product-specific findings for recommendations 10–14, 17 and 21–23.)

### Degree of Implementation Across WHO Recommendations

**Table 3** presents a consolidated view of how each country aligned with the WHO’s 24 self-care recommendations using a RAG traffic light system. This matrix was developed using triangulated data from policy review, electronic survey responses and qualitative interviews. Overall, Kenya demonstrated the greatest breadth of green-coded implementation, despite the absence of a single unifying national guideline. Uganda showed moderate alignment with a clear policy framework in place, while Nigeria’s implementation was more variable but progressing.

**Table 3:** Self-care policy mapping & implementation matrix (PMIM) showing country alignment with WHO guideline recommendations on self-care interventions (abridged)

| Category | Recommendation (abridged) |  | KENYA | NIGERIA | UGANDA |
| --- | --- | --- | --- | --- | --- |
| 1- Improving antenatal, delivery, postpartum & new-born care | Rec.1 | Health/Psych. education for women | KNGQOPC 2010 | NGN R.1 2020 | NPGSRHS 2012 |
|  | Rec.2 | Various formats for educational interventions | KNGQOPC 2010 | NGN R.2 2020 | GMN 2010 |
|  | Rec.3 | Complementary therapy for relief of nausea |  | NGN R.4 2020 |  |
|  | Rec.4 | Advice on diet & lifestyle/antacids for management of heartburn | KNGQOPC 2010 | NGN R.5 2020 | GMN 2010 & UNG 2020 |
|  | Rec.5 | Non-Rx treatment for the relief of leg cramps |  | NGN R.6 2020 |  |
|  | Rec.6 | Exercise/support/rehab for low back & pelvic pain |  | NGN R.7 2020 |  |
|  | Rec.7 | Natural fibre supplements to relieve constipation |  | NGN R.8 2020 |  |
|  | Rec.8 | Non-Rx management of varicose veins & oedema |  | NGN R.9 2020 |  |
|  | Rec.9 | Discourage pain relief during delay / augmentation in labour |  | NGN R.10 2020 |  |
| 2- High quality services for family planning & infertility services | Rec.10* | Self-administered injectable contraception | KNFPGSP 2018 | NGN R.13 2020 | UNG 2020 |
|  | Rec.11* | OTC OCPs available without a prescription | KNFPGSP 2018 | NGN R.14 2020 | UNG 2020 |
|  | Rec.12* | Home-based OPKs should be made available |  | NGN R.16 2020 | UNG 2020 |
|  | Rec.13* | Correct use of male & female condoms (STI/HIV) | KNFPGSP 2018 | NGN R.17 2020 | UNG 2020 |
|  | Rec.14* | Condom-compatible lubricants |  | NGN R.17 2020 | UNG 2020 |
|  | Rec.15 | Provide up to one year's supply of pills |  | NGN R.15 2020 | UNG 2020 |
| 3-Eliminating unsafe abortion | Rec.16 | Encourage self-assessing eligibility for MA |  |  |  |
|  | Rec.17* | Use Mifepristone & Misoprostol without direct supervision of HCP |  |  |  |
|  | Rec.18 | Self-assessing completeness of abortion process |  | NGN R.18 2020 | UNG 2020 |
|  | Rec.19* | Use self-administering injectable contraceptives |  | NGN R.19 2020 | UNG 2020 |
|  | Rec.20* | Encourage hormonal contraception after 1<sup>st</sup> pill of MA regimen | SGMM 2018 | NGN R.20 2020 | UNG 2020 |
| 4-Combating STIs, cervical cancer & gyn. morbidities | Rec.21* | HPV self-sampling during CC screening (30-60 yr) |  | NGN R.21 2020 | UNG 2020 |
|  | Rec.22* | Self-collection of samples for STD during STI testing |  | NGN R.22 2020 | UNG 2020 |
|  | Rec.23* | HIV self-testing | OMHIVST 2017 | NGN R.24 2020 | UNG 2020 |
|  | Rec.24 | Promote SRHR for women living with HIV | GPMTCT 2012 | NGN R.25 2020 |  |

\* Denotes a recommendation that is reliant on the availability of a SRHR product.
**Key:** In the Red, Amber, Green (RAG) traffic light system employed, Red denoted very low or no integration of the self-care recommendation in extant or emergent policy documents or white papers, whereas conversely Amber denoted medium levels and Green denoted high levels of integration, contextualisation or adaptation. **Legend:** **KNGQOPC** (National Guidelines for Quality Obstetrics and Perinatal Care 2010); **KNFPGSP** (National Family Planning Guidelines for Service Providers 2018); **SGMM** (Standards & Guidelines for reducing Morbidity & Mortality from unsafe abortion); **OMHIVST** (Operational Manual for the delivery of HIV Self-Testing services 2017); **GPMTCT** (Guidelines for Prevention of Mother to Child Transmission (PMTCT) of HIV/AIDS); **NGN R** (Recommendation number in National Guidelines on Self-Care for Sexual, Reproductive and Maternal Health, Nigeria 2020); **NPGSRHS** (National Policy Guidelines for Sexual & Reproductive Health Services 2012); **GMN 2010** (Guidelines on Maternal Nutrition in Uganda 2010); **UNG 2020** (Uganda National Guideline 2020)

### Implementation by Guideline Category

#### Category 1: Improving antenatal, delivery, postpartum and newborn care

Although health education is available to some extent, implementation of specific interventions such as psychosocial couple-based programmes and childbirth workshops remains limited. Geographic disparities in service availability and challenges in health literacy and language continue to hinder equitable access. Addressing these barriers will require investment in digital health tools, community outreach strategies and integration of health promotion into antenatal care pathways.

#### Category 2: Family planning and infertility services

This category had the highest overall implementation across all three countries, with green ratings for multiple recommendations including self-administered injectables, over-the-counter (OTC) oral contraceptives and condom distribution. Widespread availability of oral contraceptives and male condoms was reported, alongside increasing uptake of DMPA-SC, particularly in Uganda. Nonetheless, respondents identified persistent stigma, limited youth-friendly services and weak commodity supply chains as critical challenges. Expanding access through pharmacies and community health workers underpinned by clear regulatory frameworks may help overcome these barriers and promote greater autonomy in contraceptive decision-making

#### Category 3: Eliminating unsafe abortion

Recommendations in this category faced the greatest legal, operational and social constraints. Medical abortion remains legally restricted or ambiguously regulated in all three countries, severely curtailing implementation of recommendations 16 and 17. In addition, stigma, provider resistance and inconsistent guidance on medical abortion methods were cited as key barriers. While both Uganda and Nigeria included some abortion-related content in their national self-care guidelines, the exclusion of key recommendations reflects political sensitivities. Incremental policy shifts such as clearer protocols for post-abortion care and expanded access to accurate information may offer a starting point for progress in this domain.

#### Category 4: Tackling STIs, cervical cancer and gynaecological morbidity

Self-sampling technologies, while recognised as promising innovations, remain underutilised. Barriers include lack of awareness among both providers and users, doubts about test accuracy and concerns about logistics and follow-up care. HIV self-testing, however, stands out as an exception, having achieved significant visibility and uptake through policy integration and community campaigns. Lessons from HIVST rollouts could be leveraged to scale up self-sampling for HPV and other STIs.

### Key Interview Findings

Interviews surfaced key barriers and enablers to self-care implementation, including gaps in detailed knowledge, access disparities, stigma, legal ambiguity and variable availability of self-testing services. Selected quotes illustrate these findings.

### Awareness of WHO self-care guideline

While general awareness of the WHO guideline was high, detailed familiarity with specific recommendations varied by professional role.

> *“The self-care guideline is new and we’ve been hearing a lot about it, but only a handful could speak to specific recommendations, It really depended on your day-to-day role. I say there’s a general awareness, but when you ask about details like self-injection protocols or STI sampling, you lose most people”*. (Public Heath consultant, Nigeria)

### Geographic, literacy and supply chain barriers

Implementation was hindered by geographical disparities, limited health literacy and language barriers. Participants noted that online educational resources and childbirth education were limited in rural areas. Self-injectable contraceptives were seen as acceptable and scalable, though resupply challenges and stigma especially among youth persisted. Oral contraceptives were widely accessed over the counter, though security issues and socioeconomic disparities limited consistent uptake.

### Ambiguity around safe medical abortion

Legal ambiguity around abortion contributed to extremely low implementation of recommendations on medical abortion. Stigma and provider resistance were cited as barriers to safe abortion care, particularly for younger women. As one respondent from Kenya lamented:

> *“It’s not like you can talk about medical abortion openly because the legal situation is so unclear and you don’t know if you’re breaking the law. Younger women especially get judged harshly. Even if you ask about other options and look outside the system, it’s quite risky if the word gets out. But we hear these stories and people so find a way to get an abortion”* (Community Pharmacist, Kenya)

### Self-sampling for STIs

Self-sampling for STIs was broadly accepted in principle but not widely available in practice. Concerns around test accuracy, confidentiality and follow-up services were common. HIV self-testing stood out as a model for wider uptake of self-testing approaches.

> *“HIV self-testing has set a strong example. If we had the same community engagement and follow-up for HPV or STI tests, uptake would be much higher. The idea of self-sampling for STIs is great, but people don’t trust the accuracy, and they worry about what happens if the test is positive”* (Programme Manager SRHR, Uganda)

### Provider Training and Task Shifting

Training for healthcare providers and community-based workers was widely acknowledged as essential for expanding the reach of self-care interventions. In Uganda, for example, over 70% of adults trained by nurses were reportedly proficient in self-injection of contraceptives. However, implementation of task shifting policies remains uneven. Respondents highlighted a lack of standardised protocols, inconsistent supervision and the need for continuous capacity building.

### Cross-Cutting Barriers and Advocacy Opportunities

Structural barriers including weak supply chains, limited digital infrastructure and rural–urban health system disparities were common across all three countries. Legal restrictions, especially in relation to abortion services, were persistent impediments to policy uptake. Additionally, gaps in consumer-facing information, financing mechanisms and service integration were identified as missed opportunities. Conversely, the development of national self-care guidelines in Nigeria and Uganda, along with the growing presence of national self-care networks, were viewed as major enablers. Advocacy strategies that align WHO recommendations with local priorities and health system goals were seen as crucial to future progress. All three countries showed moderate to good alignment with the WHO Guideline (**figure 2**).

**Figure 2:**
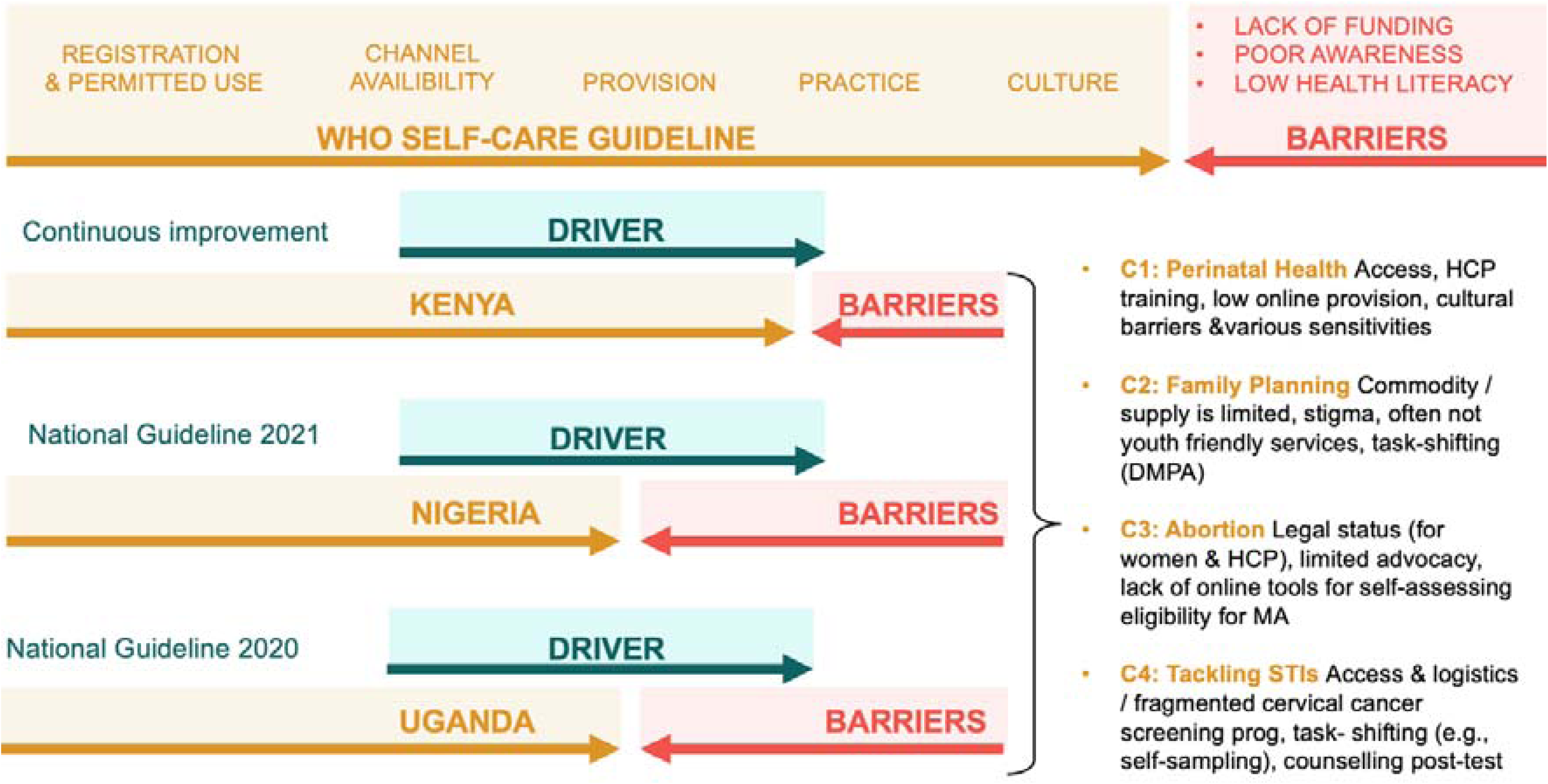
Drivers and barriers to implementation of the aspirational WHO self-care guideline recommendations in Kenya, Nigeria and Uganda

## Discussion

This study presents one of the first comprehensive assessments of how national policy environments in Kenya, Nigeria and Uganda align with the WHO Guideline on Self-Care Interventions for SRHR. By employing a mixed-methods approach and a novel Policy Mapping and Implementation Matrix (PMIM), we demonstrate how countries are variably contextualising, adapting and implementing the WHO recommendations. Our findings highlight key areas of progress, persistent implementation gaps and opportunities for targeted advocacy and system strengthening.

### Policy Adaptation and Contextualisation

The degree of alignment with the WHO Guideline varied significantly across the three countries. Kenya, in the absence of a unified national self-care policy, demonstrated relatively high levels of implementation through a constellation of sectoral documents. Nigeria and Uganda, by contrast, had formalised national guidelines that explicitly incorporated the majority of WHO recommendations 92% and 83%, respectively. This suggests that while the existence of dedicated national guidelines may support structured implementation, fragmented approaches, as in Kenya, can also yield significant progress when underpinned by enabling policies and political will. The mapping process further illustrated that alignment was strongest for recommendations related to family planning and infertility services (Category 2). Conversely, interventions associated with unsafe abortion (Category 3) and self-sampling for sexually transmitted infections (Category 4) remained poorly implemented. This is consistent with findings from other LMIC contexts where sociocultural, legal and health system constraints inhibit the operationalisation of more sensitive or resource-intensive self-care practices.

Successful implementation of self-care interventions requires a well-prepared and responsive health workforce. Respondents consistently highlighted the importance of provider training in clinical aspects of self-care (e.g., teaching self-injection techniques) and also in communication, counselling and stigma reduction. Task shifting, particularly to community health workers and pharmacists, emerged as a recurring theme. Yet its application remains uneven, limited by regulatory ambiguity and lack of standardised guidance. Strengthening national training frameworks and formalising the role of non-physician providers will be essential to expanding access and maintaining service quality.

Across all countries, implementation was impeded by broader structural and systemic issues. These included weak supply chains, insufficient funding for SRHR commodities, limited digital infrastructure and disparities in service provision between urban and rural areas. Low health literacy, gender-based discrimination and limited legal protections for reproductive rights further constrained uptake. Importantly, these barriers did not operate in isolation and often reinforced one another, creating compounded disadvantage for marginalised populations. Legal frameworks, especially those regulating abortion and adolescent access to contraception, were cited as key constraints. Inconsistent policy language, lack of alignment across ministries and politicisation of SRHR issues contributed to policy incoherence. Strengthening intersectoral governance and aligning legal, policy and programmatic instruments may help reduce these implementation bottlenecks.

### Policy and Advocacy Opportunities

By triangulating data from policies, stakeholders and practice, this study identified several actionable opportunities to strengthen the self-care policy landscape. First, the existence of control statements across a broad range of policy documents indicates latent support for self-care within existing systems. Second, where national guidelines were absent, efforts to consolidate and codify relevant provisions such as Kenya’s stakeholder-led taskforce on self-care represent an opportunity for coordinated advancement. Third, the emergence of self-care networks in all three countries has the potential to galvanise policy action and create cross-sectoral momentum (58).

Our findings also show that alignment with global guidance should not be interpreted as a goal in itself. Rather, what matters is the context-appropriate adaptation of WHO recommendations to local realities, including resource constraints, legal frameworks and cultural norms. Tailoring interventions in this way supports the WHO’s broader vision of enabling people to take greater control of their health, while also ensuring that self-care does not become a substitute for under-resourced or inaccessible health systems (59).

### Strengths and limitations

A key strength of this research is that it represents the first attempt to develop and pilot a pragmatic self-care policy mapping and implementation tool to assess how countries are contextualising the WHO Guideline on Self-Care Interventions. The comprehensive framework was applied across three diverse settings in the Global South addressing a critical gap in the literature by offering a standardised approach to evaluate alignment between national policies and global SRHR recommendations. Another major strength lies in the mixed-methods design, which integrated quantitative and qualitative data collection. The cross-sectional online survey, complemented by in-depth interviews, enabled a rich exploration of stakeholder perspectives and experiences whereas the study’s emphasis on identifying advocacy opportunities adds further value. By pinpointing control statements in national documents, the research equips self-care networks and advocates with concrete strategies to support policy and practice change. This focus on actionable recommendations enhances the study’s practical utility and potential policy impact.

The principal limitation of this study was the short study cycle (December 2020– March 2021) constrained the breadth of data collection and limited outreach to a broader respondent base. Additionally, the study did not examine consumer data on the type and volume of SRHR self-care products sold in each country, missing an opportunity to identify usage trends. We acknowledge also that the cross-sectional nature of the study provides only a snapshot in time, potentially overlooking dynamic policy developments. However, the PMIM tool was designed for periodic reapplication, enabling future longitudinal tracking of implementation progress.

Othe limitations were concerned with data collection which relied on a desk-based rapid mapping process, with COVID-19 restrictions further limiting interviews and focus groups. The online survey’s small sample size and use of convenience sampling may have introduced selection bias, affecting the representativeness of findings. Moreover, our reliance on self-reported data may have introduced social desirability and recall biases, despite the use of anonymised surveys. The study’s focus on Kenya, Nigeria and Uganda, while allowing for deep contextual analysis, may limit generalisability to countries with different sociopolitical and health system contexts. Finally, we acknowledge that because the policy assessment focused primarily on alignment with WHO recommendations, the analysis may have overlooked other critical enablers of implementation, such as resource allocation, infrastructure capacity or stakeholder engagement. Despite these limitations, this study successfully developed and tested a pragmatic methodology for rapid policy analysis, identifying both cross-cutting and context-specific drivers and barriers that can inform efforts to strengthen alignment with the WHO Guideline.

### Future work

This first iteration of the PMIM tool lays the groundwork for future policy research and advocacy. Subsequent work should aim to expand the methodology to other countries and contexts to enable benchmarking and comparative policy analysis and incorporate longitudinal tracking to monitor implementation over time. Integrating end-user perspectives, particularly among adolescents, marginalised groups and rural populations would also be important. By evaluating the effectiveness of emerging self-care networks in advancing national self-care agendas, future work should also seek to examine supply chain functionality, digital tools for self-care and financing mechanisms to support equitable access. In addition, legal and policy mapping efforts such as those led by CEHURD in Uganda offer valuable blueprints for identifying and addressing regulatory barriers to self-care implementation(60).

## Conclusion

This study offers a robust and replicable framework for assessing how countries are adapting and implementing global self-care guidance for SRHR. While significant strides have been made, particularly in the domain of family planning, persistent barriers (legal, structural and social) continue to hinder equitable access to self-care interventions. The findings highlight that success lies not in mechanical alignment with WHO recommendations, but in their contextual adaptation to national realities. As the global self-care movement gains traction, tailored policy strategies, cross-sectoral collaboration and community-led advocacy will be essential to advancing universal access to high-quality, rights-based SRHR services.

## Declarations

### Contributors

All authors provided substantial contributions to the conception (AEO, AK, RB, AM), design (AEO, AK, ERS, MK, AA), acquisition (AA, ERS, MK) and interpretation (AA, MK, ERS, RB, AM, AEO) of study data and approved the final version of the paper. AEO took the lead in planning the study with support from co-authors. AA, MK and ERS carried out the data analysis with support from the principal investigator. AEO is the guarantor.

## Funding

This research received unrestricted funding from the Population Service International. The Funder did not have a role in study design or analysis. Austen El-Osta and Azeem Majeed are supported by the National Institute for Health and Care Research (NIHR) Applied Research Collaboration (ARC) Northwest London. The views expressed are those of the authors and not necessarily those of the NHS or the NIHR or the Department of Health and Social Care.

### Consent

articipants consented to complete the survey and/or interview

### Provenance and peer review

Not commissioned; externally peer reviewed

### Data sharing statement

No additional data are available

## Data Availability

All data produced in the present study are available upon reasonable request to the authors

## Acknowledgements

The authors thank the Self-Care Trailblazer Group for disseminating the survey to country stakeholders

## Social media

Follow Austen El-Osta (@austenelosta) and The Self-Care Academic Research Unit (@ImperialSCARU) on Twitter/X.

