## Supplementary Table 1 for "Contextual Adaptation and Implementation of WHO Guideline on Self-Care Interventions for SRH in Kenya, Nigeria and Uganda"

#### KENYA TABLE OF ASSETS (n=8)

| | Title | Description |
|---|---|---|
| 1 | **National Cancer Screening Guidelines (59)** | The 2018 National Cancer Screening Guidelines were developed through a multi-stakeholder consultative process involving national, county & civil society experts. They address breast, cervical, colorectal, oral, oesophageal, prostate & childhood cancers, along with tumour markers. Regarding Cx cancer, HPV testing is recommended for women over the age of 30 years. HPV sample collection can be done by the client (self-collected) or by a health provider according to the manufacturers' instructions. |
| 2 | **HIV Self-Testing. An operational manual for the delivery of HIV Self-Testing services in Kenya (60)** | The Kenyan HIVST strategy is guided by the principles of HIV testing services as outlined in the National HTS guidelines 2015 and Guidelines for Use of Antiretroviral Drugs for Treating and Preventing HIV Infections in Kenya 2016. These guidelines outline the programmatic approaches to HIV self-testing, describe the package of support services required under HIVST, describe commodity management system requirements and outline coordination mechanisms for HIVST. They also outline quality assurance strategies, and monitoring and evaluation for HIVST. |
| 3 | **Guidelines for Prevention of Mother to Child Transmission (PMTCT) of HIV/AIDS in Kenya, 4th Edition (61)** | Guidelines are an important part of the Government strategy to reduce MTCT and is in line with the National Health Sector Strategic Plan II (NHSSPII) and Kenya National AIDS Strategic Plan (KNASP III) 2009-2013 which focuses on priority areas of prevention of new infections, improving quality of life of those infected and affected, and mitigation of social and economic impact of the infection.<br>Empowerment of WLWH is a key element of PCTCT. |
| 4 | **National Family Planning Guidelines for Service providers, 6th edition (62)** | The National Family Planning Program is mandated to provide policy direction and guidance on the implementation of FP services across the country. It is also mandated to ensure quality of FP services, capacity building and providing technical assistance to counties in the delivery of quality family planning services. These national family planning guidelines are to be used by all health services providers offering family planning services within the country. They offer guidance in the provision of modern methods of family planning in response to local challenges, as well as highlighting opportunities for increasing uptake of family planning services. |
| 5 | **National Guidelines for Quality Obstetrics and Perinatal Care (63)** | The development of this reference manual was in response to the need for emerging, updated evidence-based interventions that have proved successful when applied throughout the continuum of care of the woman's preconception, pregnancy, childbirth and the postpartum period. Both the obstetrical and medical conditions and the complications that would affect a woman during this period have extensively been described along with the management of the same. This document is designed to equip all Health care providers with maternal healthcare knowledge, skills and positive attitudes at all levels of service delivery implementation. |
| 6 | **STANDARDS and GUIDELINES for reducing morbidity & mortality from unsafe abortion in Kenya (64)** | The Ministry believes that the problem of unsafe abortion is multifaceted – it has legal, religious, gender, rights and public health dimensions among others. As such, multi-sectorial involvement is important in solving it because each sector has something special to offer. In the area of preventing unwanted pregnancies, the role of families, religious institutions, the school system, providers of contraceptives and the community as a whole must be appreciated. Once a woman has an unplanned, risky or unwanted pregnancy, professional non-judgmental counselling and provision of safe options including psychosocial support and adoption services must be provided to stop the woman from seeking unsafe abortion. For women having complications of abortion, a well-designed response system should be in place to prevent permanent disability and death. The core role and guiding principle of this document is to bring together all the aspects of care in preventing unsafe abortion using the multi-sectorial approach. |
| 7 | **NATIONAL ROAD MAP for accelerating the attainment of the MDGS related to Maternal Health and New-born Health in Kenya (65)** | The goal of the National MNH Road Map is to accelerate the reduction of maternal and new-born morbidity and mortality towards the achievement of the MDGs. The specific objectives are: 1. to increase the availability, accessibility, and utilisation of skilled attendance during pregnancy, childbirth and the post-partum period at all levels of health care delivery system; 2. to strengthen the capacity of individuals, families, communities and social networks to improve maternal and new-born health. 3. to strengthen data management and utilisation for improved MNH. |
| 8 | **NATIONAL GUIDELINES ON ESSENTIAL NEWBORN CARE (66)** | This training guide covers the following topics: 1. Universal standard precautions for infection prevention 2. Communication skills 3. Immediate care of the new-born. |

### NIGERIA TABLE OF ASSETS (n=11)

| | Title | Description |
|---|---|---|
| 1 | **Accelerated reduction of maternal and neonatal mortality in Nigeria:<br>The road map for action 2019-2021(67)** | The Roadmap aims to pick out the critical actions needed within and beyond the health sector to build momentum around the implementation of these strategies and to strengthen accountability for action among policy makers, funders, community and religious leaders and others. The Roadmap sets out nine critical areas for rapid acceleration, each with priority actions and accountable stakeholders. |
| 2 | **Antenatal care:<br>An Orientation Package for Health Care Providers<br>FMOH 2017 ANC MODEL(68)** | This orientation Package is a 1–2-day training sessions to highlight key useful, practical points for health care providers/skilled birth attendants (Physicians, Midwives, Nurses) on antenatal care. |
| 3 | **National Guidelines for the Introduction and Scale-up of DMPA-SC Self-injection(69)** | This Guideline lays out the various considerations and decisions needed to implement and scale-up a self-injection plan, including: (1) DMPA-SC Self-Injection Roll-out; (2) Provider training; (3) Client training; (4) Supply chain; (5) Waste disposal; (6) Demand generation/advocacy; (7) SI provider supervision, client follow-up, data reporting & (8) Monitoring, reporting and supervision. |
| 4 | **Family Planning Services – National Family Planning Communication Plan, Strategy for Increasing the use of Modern Contraceptives in Nigeria (2017-2020) (70)** | The National Family Planning Communication Plan (NFPCP) provides a national framework to guide FP communication interventions at all levels in Nigeria. |
| 5 | **HIV Self-Testing. Operational Guidelines for the Delivery of HIV Self-Testing in Nigeria(71)** | The guidelines outline the guiding principles, the delivery approaches & packages, demand creation & sensitization, special considerations, commodity management, human resources & training, quality assurance, monitoring & evaluation5Cs (consent, confidentiality, counselling, correct test results, connection with prevention, treatment, care & support), coordination. |
| 6 | **Comparison between self-sampling and provider collected samples for Human Papillomavirus (HPV) Deoxyribonucleic acid (DNA) testing in a Nigerian facility(72)** | The aim of the study was to assess the degree of agreement between self-sampling for HPV DNA with samples collected by a health provider. Each respondent selected from women presenting for cervical cancer screening underwent both self- and provider sampling for HPV DNA testing. This study shows moderate correlation between both sampling techniques |
| 7 | **Family Planning On-The-Job Training Manual, Facilitator's Manual(73)** | FMOH & stakeholders developed 3 OJT Training Manuals in 2012, for the 3 core areas in FP service delivery (FP Counselling, Clinical Service Provision & Contraceptive Logistics Management System (CLMS)). These documents have been revised & updated to reflect current global best practices & merged into one. **Module 1:** Counselling; **Module 2:** Service Delivery includes training on different methods available, administration of DMPA-SC, taking samples for STIs, HIV & HPV; **Module 3:** Contraceptive Logistics Management System |
| 8 | **Manual for training doctors and nurse/midwives on long-acting reversible contraceptive (LARC) methods (IUDs & Contraceptive Implants) (74)** | Training manual providing participants (service providers) with the management skills necessary to provide quality IUD and Implant services. |
| 9 | **National Health Promotion Policy(75)** | This document entails general health policy, rather than associated with WHO guidelines & their categories & discusses communicable and Non-Communicable Diseases, health literacy, poor sanitation etc., |
| 10 | **Violence Against Persons (Prohibition) Act(76)** | Describes the penalties for different crimes & violence. |
| 11 | **National strategic framework for the elimination of obstetric fistula in Nigeria 2019-2023(77)** | This document talks about Obstetric Fistula (OF) as a major public health problem in Nigeria. This National Strategic Plan (2019 -2023) provides a road map for intensification of efforts towards realising a fistula-free Nigeria by 2023. |

### UGANDA TABLE OF ASSETS (n=26)

| | Title | Description |
|---|---|---|
| 1 | **Human rights and legal dimensions of self-care interventions for sexual and reproductive health(78)** | Broad article on Human rights & legal dimensions of self-care interventions for sexual and reproductive health, but not specific to Uganda. |
| 2 | **Essential Medicines and Health Supplies List for Uganda (EMHSLU), Pharmacy Division(79)** | The Essential Medicines and Health Supplies List for Uganda (EMHSLU) 2016 is a list of safe, efficacious, and cost-effective medicines and health supplies that suit the health care needs of the majority of the Ugandan population. |
| 3 | **National Policy Guidelines for Sexual and Reproductive Health services, Reproductive Health division, Department of Community Health(80)** | National Self-Care Guidelines for SRHR, with a focus on STIs, Family Planning, Antenatal Care & Prevention and Management of unsafe abortions |
| 4 | **Lesson 10 How to counsel clients on self-injection, PATH/Therese Bjorn Mason(81)** | Training material outlining Part of the Subcutaneous DMPA (DMPA-SC) training for health workers provided by PATH/Therese Bjorn Mason |
| 5 | **Integrated cervical cancer screening in Mayuge District Uganda (ASPIRE Mayuge): a pragmatic sequential cluster randomized trial protocol(82)** | This study compares the effectiveness of two cervical cancer screening models for self-collected HPV testing: 1) community health worker recruitment (door-to-door); and 2) community health meetings. Results from this study will inform the national scale-up of cervical cancer screening in Uganda |
| 6 | **Engaging a Key Population Drop-In Centre to Increase Uptake of HIV Self-Testing Services: The Experience of Busia Health Centre IV in East Central Uganda(83)** | Case study: team carried out interventions (HCW training, providing HIV-ST tools) to increase uptake of HIV-ST services in East Central Uganda. |
| 7 | **HIV Self testing among Key populations and sexual partners of new mothers in Uganda(84)** | Authors present the preliminary findings of a HIVST program in Kampala, Uganda. Concluded that HIV self-testing is a promising intervention, though confirmatory HIV testing and linkage to care remains a challenge. There is a need to strengthen community confirmatory testing. |
| 8 | **HIV Self-Test Targeted Distribution and Linkage Plan Summary(85)** | Global evidence for use of HIV self-test (HIVST) & strategy for apply to Uganda's target population |
| 9 | **Facing Uganda's law on abortion - Experiences from Women & Service Providers(86)** | National Policy Guidelines for Sexual and Reproductive Health Services 2012 provides direction and focus on provision of reproductive health services and clarify the roles of the different actors involved in planning, implementation, service provision, monitoring and evaluation. |
| 10 | **Uganda Condom Market Champion Scope of Work(87)** | Comprehensive Condom Programming Strategy 2020–2025. [does not have date or author] |
| 11 | **Uganda Clinical Guidelines 2016 – National Guidelines for Management of Common Conditions(88)** | This document listed 1100+ pages of common illnesses, but only general information on how it is contracted, symptoms & how to treat. |
| 12 | **Addendum to the HIV Testing Services Policy & Implementation Guidelines(89)** | Policy & Guidelines on HIV self-testing, describing how implementation can be carried out in Uganda |
| 13 | **Theory of Self-injection Scale up(90)** | Outline's activities and indicators for service expansion for self-injection of DMPA-SC. |
| 14 | **Strategic Plan for Cervical Cancer Prevention and Control in Uganda, 2010–2014(91)** | This strategic plan describes the method of HPV testing (DNA testing). There is no information on self-sampling. This document is no longer up to date & accurate. |
| 15 | **Uganda National Self-care guidelines for Sexual & Reproductive health & Rights(92)** | In 2019, WHO published the consolidated guideline on Self-Care interventions for health focusing on SRHR. The Uganda Self-Care Expert Working Group having reviewed all the 24 recommendations and based on the situational analysis findings adopted a phased approach to adaptation and implementation of the recommendation. |
| 16 | **HIV self-testing & HIV assisted partner notification services policy & implementation guidelines(93)** | Policy statement: "HIV self-testing shall be offered as an additional approach to HIV testing services in Uganda." Guide on how HIVST services should be integrated into existing community-based and facility-based HTS approaches and tailored to specific sub-populations. |
| 17 | **Adolescent Health Policy Guidelines & Service Standards(94)** | Policy guidelines outlining health policy guidelines for adolescents. |
| 18 | **How to take your own sample for a HPV test(95)** | Pamphlet outlining HPV self-sampling instructions. Provided only by a healthcare provider during a consultation. |
| 19 | **Quick Reference Guide Self-Collected Vaginal Sample for HPV Test(96)** | Fact sheet discussing the new Cervical Screening Test and pathway. Detailing: Who is eligible for self-collection? Who is not eligible for self-collection? Supporting patients in clinical management, & Requesting pathology tests for Self-collect. |
| 20 | **CONSTITUTION OF THE REPUBLIC OF UGANDA(97)** | The constitution of the Republic of Uganda highlighting the National Objectives and Directive Principles of State Policy |

| | | |
|---|---|---|
| **21** | **DMPA-SC Self-injection virtual TOT(98)** | This training is intended for individuals who will be facilitating health worker training and supervision for DMPA-SC self-injection. PPT are expected to have completed previous training on how to administer and provide counselling on DMPA-SC. |
| **22** | **Module 7 HPV Detection Tests & Sample Collection Techniques for HPV Testing(99)** | HPV sample collection training program for HCWs. |
| **23** | **DMPA-SC Self-injection virtual training for partners(100)** | DMPA-SC training program for PATH partners. |
| **24** | **The Penal Code Act(101)** | Ugandan Criminal Code. Including laws for attempts to procure abortion & procuring miscarriage. |
| **25** | **Provision of DMPA-SC Self-injection Circular(102)** | Circular from Ministry of Health stating: "All stakeholders are requested to take note of self-injection approval in Uganda, and this takes effect immediately. All health workers who train women to self-inject are supposed to use the approved guidelines on self-injection by the Ministry of Health." |
| **26** | **Proposed Self-injection guidelines for Uganda(103)** | PATH guidelines for self-injection in Uganda. |
| **27** | **Observation checklist to assess health workers orienting clients on DMPA-SC self-injection(104)** | Checklist to assess health workers orienting clients on DMPA-SC. |
| **28** | **The National Policy Guidelines and Service Standards for Sexual and Reproductive Health and Rights(105)** | This document has two sets of guidelines: service policy guidelines and service standards that aim at making explicit the direction of reproductive health and how services should be provided at all levels of health care. The document is to provide explicit direction and focus on provision of reproductive health services as well as guide the implementation of a coherent and coordinated reproductive health programme. |
