## Supplementary Table 2 for "Contextual Adaptation and Implementation of WHO Guideline on Self-Care Interventions for SRH in Kenya, Nigeria and Uganda"

### 3.1 CATEGORY ONE

#### IMPROVING ANTENATAL, DELIVERY, POSTPARTUM & NEW-BORN CARE (Recommendations 1-9)

##### Table 6: Summary of main research findings for recommendations 1-9 (category one)

| | |
|---|---|
| **R.1**<br>**Health education for women**<br><br>**(1a):** Childbirth training workshops<br>**(1b):** Nurse-led applied relaxation training program<br>**(1c):** Psychosocial couple-based prevention program<br>**1d:** Psychoeducation | **Desktop research**<br>• Difference in utilization of CS across different settings (rural-urban, regional & socioeconomic divide) suggesting unmet needs and inequitable access to CS [38]. Some online services & teaching materials targeting healthcare workers, pregnant women and their partners are available (1) but in general, there is a lack of information/data on health education. Educating women only once during their first antenatal care visit, minimal participation by women in the education session & lack of evidence for planning for sessions shows there is a need to raise awareness amongst HCW & the public (2).<br>**Sentiment analysis**<br>• Childbirth training workshops, nurse-led applied relaxation training program, psychosocial couple-based prevention program and psychoeducation are available to some extent in all three countries.<br>• There is a need to implement differential levels of interventions across districts to improve availability and address inequalities in access.<br>**Key barriers**<br>• Key barriers include high Illiteracy levels, limited time for teaching.<br>**Key drivers**<br>• WHO Rec 1 is covered in Nigeria's National Guidelines on Self-care for SRHR NREC 1. |
| **R.2**<br>**Provide information in various formats** | **Desktop research**<br>• Research in Kenya shows available Infographics, but general awareness may be low. The format of educational programs matters to these women and group teachings were reported to be the most favourable due to convenience [53]. In general, gaps in the evidence base on the availability of information in various formats in all three countries.<br>**Sentiment analysis**<br>• The majority of the survey respondents agree that information is available in various formats to some extent in all three counties.<br>**Key barriers**<br>• Various languages spoken per country, and some digital exclusion.<br>**Key drivers**<br>• WHO Rec 2 is covered in Nigeria's National Guidelines on Self-care for SRHR NREC 2. |
| **R.3**<br>**Complementary therapy for relief of nausea** | **Desktop research**<br>• Drowsiness is a common side-effect of various antihistamines used to treat nausea and vomiting and pharmacological options could be expensive and not readily available.<br>• There is an evidence gap in the availability/promotion of ginger, chamomile, vitamin B6 and acupuncture to relieve nausea in all three countries.<br>**Sentiment analysis**<br>• Non-pharmacological options to relieve nausea in early pregnancy are cheap & effective, and recommended to some extent in all three countries.<br>**Key barriers**<br>• Lack of provision/availability, lack of data<br>**Key drivers**<br>• WHO Rec 3 is covered in Nigeria's National Guidelines on Self-care for SRHR NREC 4. |
| **R.4**<br>**Advice on diet & lifestyle/antacids for heartburn** | **Desktop research**<br>• Prevents women from having a positive pregnancy experience. Antacids may impair other drugs' absorption and should not be taken within two hours of iron and folic acid supplements.<br>• Uganda's National Guidelines on Maternal Nutrition 2010 recommends dietary modification, eat small frequent meals, adequate chewing and eating slowly, eating at least three hours before going to bed etc., to address heartburn during pregnancy(3). Uganda National SC guidelines for SRHR (2020) recommends Self-care management of heartburn using Magnesium Trisilicate.<br>• National Guidelines for Quality Obstetrics and Perinatal Care 2010 Kenya**:** provides Practical Considerations/tips like dietary modification, slow eating, and chew food well, avoid lying down for at least one to two hours after eating, elevate the head of the bed, wear loose-fitting clothing etc., for women suffering from heartburn. Recommends antacids only after consulting a physician(4).<br>**Sentiment analysis**<br>• Survey findings suggest that treatment of heartburn is addressed in Nigeria and Uganda but only to some extent in Kenya.<br>**Key barriers**<br>• None identified<br>**Key drivers**<br>• WHO Rec 4 is covered in Nigeria's National Guidelines on Self-care for SRHR NREC 5. |

<table>
<tr><td></td><td><ul><li>WHO Rec 4 is covered in Kenya’s National Guidelines for Quality Obstetrics and Perinatal Care 2010.</li><li>WHO Rec 4 is covered in Uganda’s Guidelines on Maternal Nutrition 2010 and National Guideline for SRHR 2020.</li></ul></td></tr>
<tr><td>R.5<br>Non-Rx treatment for the relief of leg cramps</td><td>Desktop research<ul><li>For pregnant women, leg cramps overnight can cause sleep disorders such as sleep loss and insomnia, affecting the outcome of labour, including the length of labour and mode of delivery(5, 6). One prospective, observational study, including 131 pregnant women, found that pregnant women sleeping less than six hours per night and those with a severe sleep problem were, respectively, 4.5 times and 5.2 times more likely to undergo a caesarean delivery(6).</li><li>Leg cramps in pregnancy may also be related to depression which can increase placental corticotropin-releasing factor and initiate uterine contractions and cervical ripening, and might eventually cause labour difficulty, foetus hypoxia, and increased risks of neonatal asphyxia and postpartum haemorrhage(7-9)</li><li>Research gap on the availability non-pharmacological treatment options (magnesium, calcium etc in all three countries.</li></ul>Sentiment analysis<ul><li>Our respondents suggest pregnant women are advised to some extent about different treatment options (Magnesium, calcium, or non-pharmacological treatment options) to relieve leg cramps in all three countries.</li></ul>Key barriers: Lack of data<br>Key drivers: WHO Rec 5 is covered in Nigeria’s National Guidelines on Self-care for SRHR NREC 6.</td></tr>
<tr><td>R.6<br>Physio etc., for lower back / pelvic pain</td><td>Desktop research<ul><li>This pain can have an adverse impact on the quality of life (QOL) as it interferes with work, daily activities and affected women's sleep. Women reporting PP were less mobile than those reporting LBP only and experienced more co-morbidity and depressive symptoms (10)</li><li>There is a research gap on interventions available for low back and pelvic pain.</li></ul>Sentiment analysis<ul><li>Respondents of all three countries agree that different treatment options (e.g., Exercise, physio, support belts, acupuncture) to help prevent lower back & pelvic pain are available.</li></ul>Key barriers<ul><li>Lack of provision/availability, lack of data</li><li>Lack of resources</li></ul>Key drivers: WHO Rec 6 is covered in Nigeria’s National Guidelines on Self-care for SRHR NewREC 7.</td></tr>
<tr><td>R.7<br>Natural fibre supplements to relieve constipation</td><td>Desktop research<ul><li>May lead to serious adverse consequences, including physiological and psychological ill-health, low self-esteem, and poor quality of life for the mother (11, 12)</li><li>Uganda’s national Guidelines on Maternal Nutrition 2010 advise pregnant women to take a diet containing plenty of fluid, fruit juice and fibre, encourage regular exercises, Discourage the use of laxatives, and advise pregnant women on constipation resulting from the use of iron supplementation(3).</li><li>There is evidence gap specifically on the use of wheat bran or other fibre supplements to relieve constipation.</li></ul>Sentiment analysis<ul><li>Respondents from all three countries suggest that pregnant women are recommended to use wheat bran or a fibre supplement to some extent to relieve constipation.</li></ul>Key barriers: None identified<br>Key drivers: WHO Rec 7 is covered in Nigeria’s National Guidelines on Self-care for SRHR NREC 8.</td></tr>
<tr><td>R.8<br>No-Rx options for management of varicose veins & oedema</td><td>Desktop research<ul><li>The symptoms associated with varicose veins are a significant cause of morbidity and have a negative impact on the quality of life (13).</li><li>Uganda’s national Guidelines on Maternal Nutrition 2010 advise pregnant women to rest with legs elevated, encourage them to lie on their side while sleeping and Consult with the health worker to rule out other causes of oedema. Discourage the use of diuretics in pregnancy and advises pregnant women not to reduce salt intake unless medically recommended.</li><li>Research gap on the use of non-pharmacological options to treat varicose veins and oedema across three countries.</li></ul>Sentiment analysis<ul><li>Survey findings suggest the availability of non-pharmacological options (comp. sock, leg elevation, immersion) for varicose veins & oedema to some extent.</li></ul>Key barriers<ul><li>Lack of resources</li></ul>Key drivers<ul><li>WHO Rec 8 is covered in Nigeria’s National Guidelines on Self-care for SRHR NREC 9.</li><li>WHO Rec 8 is covered in Uganda’s Guidelines on Maternal Nutrition 2010.</li></ul></td></tr>
<tr><td>R.9<br>Discourage pain relief for augmentation in labour</td><td>Desktop research<ul><li>Inappropriate augmentation of labour can cause harm, and unnecessary clinical intervention in the natural birth process undermines women's autonomy and dignity as recipients of care. It may negatively impact their childbirth experience(14).</li><li>The procedure carries the risk of uterine hyperstimulation, with the potential consequences of fetal distress and uterine rupture.</li><li>Induction of labour is not a common occurrence in Kenya. (15)</li><li>Research gap in the use of pain relief for augmentation in labour in all three countries.</li></ul>Sentiment analysis<ul><li>Survey findings suggest that the use of pain relief for preventing delay & reducing the use of augmentation in labour is avoided to some extent in Uganda and Kenya but is not avoided in Nigeria.</li></ul>Key barriers<ul><li>Lack of awareness</li></ul>Key drivers<ul><li>WHO Rec 9 is covered in Nigeria’s National Guidelines on Self-care for SRHR NREC 10.</li></ul></td></tr>
</table>

### 3.2 CATEGORY TWO

#### FAMILY PLANNING (Recommendations 10-15)

##### Table 9: Summary of main research findings for recommendations 10-15 (category two)

| | |
|---|---|
| **R.10**<br>**Self-administered injectable contraception** | **Desktop research**<br>• Recently, new forms of injectable contraception have been developed, which allow for subcutaneous injection (under the skin), rather than intramuscular injection (16). Some people using injectable contraception may prefer to self-inject for reasons of privacy or convenience.<br>• It has been suggested that self-administration of injectable contraception can equal or improve contraceptive continuation rates compared with provider-administration (16). This benefit comes without notable increases in pregnancy or safety concern.<br>**Sentiment analysis**<br>• In Kenya, DMPA-SC is the most used short-acting method of contraception (50%), followed by the oral contraceptive pill (28%) and condoms (19%) (17). It is widely accepted within the community (18), with reinjection rates of 89%, 81%, and 68% at 3, 6, and 9 months, respectively (19). Given this, DMPA-SC is more practical in urban setting because of the accessibility of facilities and distribution of commodities as well as the empowerment/awareness of urban residents (personal meeting).<br>• Though Nigeria has permitted the use of self-injectables, A 2012 study in Nigeria showed that more than 50% of sampled women using DMPA-IM did not continue within one year (20). However, our findings indicate that women are counselled & offered a few months' supply of self-injectables, with a variety of materials (images, written etc.,).<br>• Self-injection was granted approval by the Ugandan National Drugs Authority in 2017 (21), & Uganda is currently undergoing rapid scale up to offer DMPA-SC nationwide (22). Our data indicates that HCPs are somewhat trained to teach individuals to self-administer & dispose of injectable contraceptives. Given this, HCWs sometimes refuse to provide contraception to unmarried adolescents due to deeply rooted stigma (23), which is further exaggerated by the lack of youth-friendly services, of which non-judgmental, supportive front line health care providers are the critical component (24, 25). Through personal interviews, we gathered that HCWs feel that women do not have the capability to self-care, & there are restrictions on advertisement, stagnating public awareness of self-injectable contraceptives.<br>**Key barriers**<br>• HCWs were insufficiently trained to teach individuals to self-administer & dispose of injectable contraceptives.<br>**Key Drivers**<br>• WHO Rec 10 is covered in Nigeria's National Guidelines on Self-care for SRMH NREC 13.<br>• WHO Rec 10 is covered in Uganda's National Self-care Guidelines for SRHR UREC 10. |
| **R.11**<br>**OTC oral contraceptive pills are available without a prescription** | **Desktop research**<br>• Oral contraceptives (OC), both combined oral contraceptives (COC) & progestogen-only pills (POP), are widely used effective methods of birth control. However, access to OCs varies globally—in some countries, OCs are available over the counter (OTC), while other countries restrict access to OCs either by requiring eligibility screening by trained pharmacy staff before dispensation (pharmacy access, or behind-the-counter availability), or by requiring a healthcare provider's prescription (26).<br>**Sentiment analysis**<br>• Oral contraceptive pills are available OTC in Kenya (27). We found that there are policies in place to promote the use of OTC oral contraceptive pills & we HCPs are trained to recommend & prescribe OTC oral contraceptive pills. Personal interviews with stakeholders revealed that the availability of oral contraceptive pills varies (more available in urban compared to rural setting), and there exists a lack of women's empowerment in rural regions.<br>• We found that oral contraceptive pills are available OTC in Nigeria, policies are in place to promote the use of OTC oral contraceptive pills, & HCPs are trained to promote & prescribe pills.<br>• Oral contraceptive pills are available OTC in Uganda, through screening is required in Uganda (28). Policies are in place to promote the use of OTC oral contraceptive pills, and our respondents indicated that HCPs are trained to promote & prescribe OTC pills. Through personal meetings we gathered that there lies friction in the interaction between HCPs & public which should be reduced, & availability is scarcer in rural settings.<br>**Key barriers**<br>• Low level of awareness within community, fear of stigma.<br>**Key Drivers**<br>• WHO Rec 11 is covered in Nigeria's National Guidelines on Self-care for SRMH NREC 14.<br>• WHO Rec 11 is covered in Uganda's National Self-care Guidelines for SRHR UREC 11. |
| **R.12**<br>**Home-based OPKs should be made available** | **Desktop research**<br>• Infertility is a huge problem globally. Global estimates show that approximately 15% - 25% of couples are unable to become pregnant despite attempting for five years or more (29).<br>• OPKs predict the surge of luteinising hormone (LH) that precedes ovulation while also tracking corresponding oestrogen levels. OPKs do not directly pinpoint a peak fertility day & may need multiple pregnancy attempts within the appropriate timeframe during a woman's menstrual cycle (30).<br>**Sentiment analysis** |

| | |
|---|---|
| | • Our respondents indicated that there are no policies in place & staff are inadequately trained to offer individuals advice on the use of home-based OPKs in Kenya. The main obstacles to the uptake of OPKs in Kenya, according to personal interviews, are the lack of government support, & the somewhat consequential lack of awareness of OPKs in community setting.<br>• Policies are in place which promote the use of home-based OPKs, though our respondents gave mixed opinions on whether Nigerian HCWs were adequately trained to offer advice on OPKs. Strong public awareness of OPKs in Nigeria, possibly through word of mouth or through posters in pharmacies (personal meeting).<br>• There are no policies in place which promote the use of home-based OPKs, and our respondents confirmed that Ugandan HCWs are not trained to offer advice on the use of OPKs. Public awareness amongst the Ugandan public is low, as a result, the prevailing option is to see a doctor/GP (personal meeting).<br>**Key barriers**<br>• Generally, there is a need to raise awareness about SRHR benefits of OPKs using accessible media / infographics, radio etc.,<br>**Key Drivers**<br>• WHO Rec 12 is covered in Nigeria's National Guidelines on Self-care for SRMH NREC 15.<br>• WHO Rec 12 is covered in Uganda's National Self-care Guidelines for SRHR UREC 12. |
| **R.13<br>Correct use of male & female condoms** | **Desktop research**<br>• Condoms are a critical component in a comprehensive & sustainable approach to the prevention of HIV & other sexually transmitted infections (STIs) and are effective for preventing unintended pregnancies (31).<br>• Male & female condoms are the only devices that both reduce the transmission of HIV and other sexually transmitted infections (STIs) & prevent unintended pregnancy (31).<br>**Sentiment analysis**<br>• Condom use appears to be low amongst Kenyans (32), & reports from a survey found that 20% of individuals incorrectly used condoms (33). Our data indicates that there is a good understanding in the general population of the use of condoms for the prevention of STDs, & they are widely available free of charge.<br>• Condoms are widely available in Nigeria (34), and 100% of our respondents suggested that there is a good understanding of the benefits of condom use within the community setting. In a deeply religious country where many Roman Catholics & Muslims oppose contraception; politicians & doctors broach the topic gingerly, and change is slow. Posters promote 'birth spacing,' not 'birth control.' Supplies of contraceptives are often erratic (35). Our data indicates that condoms not necessarily always free of charge in Nigeria.<br>• Though condoms are widely available in different settings in Uganda (36), they are more scarcely available in rural settings, compared to urban settings, resulting in lower usage of condoms in this setting (37). Furthermore, increasing condom prices are becoming an issue in Uganda (36). Our respondents indicated that there is generally a good understanding of the STD preventing benefits of condoms, and condoms are occasionally offered free of charge.<br>**Key barriers**<br>• Quality control remails an issue.<br>**Key Drivers**<br>• WHO Rec 13 is covered in Nigeria's National Guidelines on Self-care for SRMH NREC 16.<br>• WHO Rec 13 is covered in Uganda's National Self-care Guidelines for SRHR UREC 13. |
| **R.14<br>Correct use of condom-compatible lubricants** | **Desktop research**<br>• Strategic partnerships among all main partners are essential to improve access to, and use of, condoms to prevent or reduce the incidence of unintended pregnancies, STIs & HIV (38).<br>• A study found that individuals who always used condoms correctly were almost 60% less likely to be diagnosed with an STI (39).<br>**Sentiment analysis**<br>• We found that condom compatible lubricants were somewhat available & accessible in Kenya.<br>• Though condom compatible lubricants are somewhat available & accessible in Nigeria, the Director-General, Standards Organisation of Nigeria, Osita Aboloma, has disclosed that over 70% of the lubricants in the market failed the quality parameters of the Nigerian Industrial Standard (40).<br>• Condom lubricants accessibility & availability appear to be scarce in Uganda. This is an area of concern, since there is an association between inconsistent condom use & low condom efficacy (41).<br>**Key barriers**<br>• Generally, condom-compatible lubricants may be overlooked; counselling & instructions are required to accompany printed instructions.<br>**Key Drivers**<br>• WHO Rec 14 is covered in Nigeria's National Guidelines on Self-care for SRMH NREC 17.<br>• Who Rec 14 is covered in Uganda's National Self-care Guidelines for SRHR UREC 14. |
| **R.15<br>Provide women with a year's supply of POP & COC pill packs** | **Desktop research**<br>• Oral contraceptives (OC), both combined oral contraceptives (COC) & progestogen-only pills (POP), are widely used effective methods of birth control. However, access to OCs varies globally—in some countries, OCs are available over the counter (OTC), while other countries restrict access to OCs either by requiring eligibility screening by trained pharmacy staff before dispensation (pharmacy access, or behind-the-counter availability), or by requiring a healthcare provider's prescription (26).<br>**Sentiment analysis**<br>• Our respondents indicated that women are often not provided with up to a year's supply of pulls across all three countries.<br>**Key barriers**<br>• Public awareness of COC & POP is generally low.<br>**Key Drivers**<br>• No data found |

### 3.3 CATEGORY THREE

#### ELIMINATING UNSAFE ABORTION (Recommendations 16-20)

**Table 12: Summary of main research findings for recommendations 16-20 (category three)**

| | |
|---|---|
| **R16: Encourage self-assessing eligibility for MA** | **Desktop research**<br>• Self-assessment & self-management approaches can be empowering for individuals, helps triage care by optimizing available health workforce resources & sharing of tasks & reduces the need for unsafe abortions & the complications that they bring.<br>• Self-assessment requires knowledge or access to knowledge about contraceptives, pregnancy & abortions, however this topic is taboo therefore it is not openly talked about.<br>**Sentiment analysis**<br>• Ministry of Health in Uganda is largely led by religious views within strong traditional cultural & value system. This topic is also problematic in a catholic dominated region<br>• Advocacy for abortion remains very fragile and limited.<br>• Some very strong religious groups, like the Evangelist acting against<br>**Key barriers**<br>• All three countries, like many African countries, do not allow abortions unless women meet a list of conditions, however unplanned pregnancies occur frequently, forcing women to make difficult decisions.<br>• There is a common theme of a lack of clarity & transparency regarding the status of abortion law in both Kenya & Nigeria, causing secrecy around the subject<br>**Key Drivers**<br>• WHO Rec 16 is discussed in Nigeria's National Guidelines on Self-care but is not included in their list of SRMH NRECs. No national guidelines for Kenya & Uganda |
| **R17: Use Mif & Mis without direct supervision of HCP** | **Desktop research**<br>• Medical management of abortion generally involves either a combination regimen of mifepristone & misoprostol, or a misoprostol-only regimen<br>• Self-managed abortion process would allow women to discretely & privately deal with their abortions.<br>• Mif & Mis in Uganda requires a prescription; however, it appears this is not consistent throughout the country as many pharmacists will sell them regardless. The quality of the products also has been reported to be low, often where packaging instructions are not included or are inaccurate.<br>• Nigeria has a large unregulated drug market, where drug sellers sell medications, including abortion pills without prescriptions. They often have poor knowledge of MA drugs, and commonly sell medications without packaging or instructions & often provide inadequate or inaccurate information to women about medications, side effects & potential complications (42).<br>**Sentiment analysis**<br>• No interviews conducted on this recommendation<br>**Key barriers**<br>• Products available appear to be low, often where packaging instructions are not included or are inaccurate<br>• There have been reports of inconsistencies with the requirement of prescriptions<br>**Key Drivers**<br>• WHO Rec 17 is discussed in Nigeria's National Guidelines on Self-care but is not included in their list of SRMH NRECs. No national guidelines for Kenya & Uganda |
| **R18: Self-assessing completeness of abortion process** | **Desktop research**<br>• Assessing the status of a pregnancy is a crucial step of post-abortion care (PAC) after taking abortion pills in order to check the process was complete<br>• Women who do not check the status of their pregnancy are at a risk of incomplete abortions which can be fatal, cause permanent damage or be at risk of secondary infertility<br>• Pregnancy tests available but supply, cost vary & awareness in relation to self-assessment variable<br>**Sentiment analysis**<br>• The Kenyan government supports the use of pregnancy tests in the context of family planning services through the Division for Reproductive Health's National Family Planning Guidelines<br>**Key barriers**<br>• There needs to be more research on this aspect of PAC as there is not much information or policies about using pregnancy tests post-abortion<br>• Lack of availability in public sector clinics<br>**Key Drivers**<br>• WHO Rec 18 is covered in Nigeria's National Guidelines on Self-care for SRMH NREC 18. This recommendation is also mentioned in Kenya's Standards & Guidelines for reducing morbidity & mortality from unsafe abortions as an important measure of PAC. No national guidelines for Uganda. |

| | |
|---|---|
| **R19: Use self-administering injectable contraceptives in specific circumstances** | **Desktop research**<br>• Providing post-abortion care (PAC) by the provision of contraceptive services reduces the likelihood of repeat abortions (43, 44)<br>• This short-acting method of contraception is an attractive option for women due to its ease of use & acceptance in the community.<br>• In order to reduce the recurrence of abortions, post-abortion care service providers need to provide long-acting methods contraceptive counselling, family planning and effective pregnancy prevention methods before discharge from the health care facility (45)<br>**Sentiment analysis**<br>• More funding is needed to support uptake of guidelines which will make the supplies available<br>• It is more practical in urban setting because of the accessibility of facilities and distribution of commodities as well as the empowerment /awareness of urban residents, suggesting rural areas may have issues with accessibility<br>• There is a policy in place in Uganda for DMPA-SC, however it has not been implemented yet as this is a fairly new recommendation. Self-injectables are also mostly available in the private setting & there are restrictions on advertising<br>**Key barriers**<br>• Post-abortion care (PAC) needs more research and dedicated policies, especially including appropriate contraceptives.<br>• Funding for supplies<br>**Key Drivers**<br>• WHO Rec 19 is covered in Nigeria's National Guidelines on Self-care for SRMH NREC 19. PAC is also mentioned in Kenya's Standards & Guidelines for reducing morbidity & mortality from unsafe abortions. In Part 3. No national guidelines for Uganda. |
| **R20: Encourage hormonal contraception immediately after 1st pill of MA regimen** | **Desktop research**<br>• The WHO Guidelines recommends that contraception should be initiated at the time of administration of the first pill of the medical abortion regimen or after assessment of successful medical abortion.<br>• This is an important part of post-abortion care (PAC) as this is an opportunity for women to gain better understanding of the contraceptives that could suit their lifestyle, thus empowering themselves to have safe sex.<br>• Developing a good relationship with contraceptives will significantly reduce the likelihood of needing an abortion – therefore, there should be better policies to embed family planning into the community<br>**Sentiment analysis**<br>• No interviews conducted on this recommendation<br>**Key barriers**<br>• More research needs to be done on this topic<br>**Key Drivers**<br>• WHO Rec 20 is covered in Nigeria's National Guidelines on Self-care for SRMH NREC 20. PAC is mentioned in Kenya's Standards & Guidelines for reducing morbidity & mortality from unsafe abortions in Part 3. No national guidelines for Uganda. |

### 3.4 CATEGORY FOUR

#### COMBATING STIs & SELF-SAMPLING (Recommendations 21-24)

STIs have a profound impact on SRHR worldwide as more than 1 million STIs are acquired every day. In 2016, WHO estimated 376 million new infections with 1 of 4 STIs: chlamydia (127million), gonorrhoea (87 million), syphilis (6.3 million) & trichomoniasis (156 million). STIs can have serious consequences beyond the immediate impact of the infection itself including an increased risk of HIV infection, infertility, foetal or neonatal death (46). In 2016, 988 000 pregnant women were infected with syphilis, resulting in over 350000 adverse birth outcomes including 200000 stillbirths and new-born deaths (47). In LMICs, diagnostic tests remain largely unavailable. Where testing is available, it is often expensive & geographically inaccessible. Other barriers include: limited resources, stigmatization and poor quality of services (48). The only inexpensive, rapid tests currently available for STIs are for syphilis & HIV. Here again, self-sampling would provide a much-needed alternative to provider-driven sampling and an additional option for service users. Over two-thirds of all people living with HIV (25.7 million) live in the WHO African Region (49). HIV self-testing offers an additional option for testing, especially for the marginalised & most at-risk groups. However, no single test can provide a full HIV diagnosis; confirmatory testing is required and must be conducted by a qualified & trained health or community worker at a community centre or clinic, hence the need for efficient and accessible follow-up structures and resources, in addition to the kits themselves(49). Two HPV types (16 & 18) are responsible for 70% of cervical cancers & pre-cancerous cervical lesions. Cervical cancer is the 4th most common cancer among women globally, and nearly 90% of the 311,000 deaths worldwide in 2018 occurred in LMICs (50). HPV vaccination does not replace cervical cancer screening (50).

##### Table 15: Summary of main research findings for recommendations 21-24 (category four)

| | |
|---|---|
| **REC 21 (NEW): HPV self-sampling for cervical cancer screening (30–60 years)** | **Desktop research**<br>• Cervical cancer remains one of the most common cancers among women in all 3 countries<br>• Self-sampling/ collection has not been reported to be widely implemented yet<br>• Local case studies conducted on the acceptability of self-sampling reveal it would be a favourable option for most women<br>**Sentiment analysis**<br>• Visible in policy documents regarding cervical cancer screening, but still in pilot phase.<br>• Mostly unavailable in Nigeria & Uganda, only partially available in Kenya<br>**Key barriers**<br>• The chronic shortage of healthcare workers in low-resource settings tends to be a barrier to scale-up and training of HCPs (51)<br>• Lack of awareness and financing<br>• Absent or limited HPV national screening plans<br>**Key Drivers**<br>• Recommended as an additional screening tool in National Policy documents in all 3 countries<br>• Studies show high acceptability, reliability & feasibility |
| **REC 22 (NEW): Self-collection of samples for 4 STIs**<br><br>(22a) Neisseria gonorrhoeae & Chlamydia trachomatis<br><br>(22b) Treponema pallidum (syphilis) & Trichomonas vaginalis | **Desktop research**<br>• These 4 STIs are implicated in increasing the risk of HIV acquisition & transmission. Moreover, people with STIs often experience stigma, stereotyping, vulnerability, shame and gender-based violence. (52)<br>• Data shows it can increase overall uptake of STI testing services, decrease future costs (though they may be borne by users), high satisfaction, comfort, ease, privacy, convenience, and confidence – especially after the experience, even though some users experienced pain, discomfort and concerns about safety. (53)<br>**Sentiment analysis**<br>• Lack of national screening policies, particularly problematic for asymptomatic cases, but started being considered following the publication of the WHO guidelines in Kenya & Nigeria<br>• Testing supposedly free in public settings and/or STI clinics<br>**Key barriers**<br>• Limited availability and awareness, both from the public and HCPs<br>• Lack of logistic structure for accessing kits, processing of samples & follow-up care<br>**Key Drivers**<br>• Addressed in Nigeria's and Kenya's National Guidelines on Self-care |
| **R23: HIV self-testing (HIVST)** | **Desktop research**<br>• Prevalence of HIV infection remains high in Kenya, Nigeria & Uganda<br>• Self-testing appears to be an acceptable & effective option, especially among at-risk groups targeted by discrimination, stigma & violence such MSM, sex workers & people who inject<br>**Sentiment analysis**<br>• HIVST already implemented in Kenya with good results<br>• Not addressed in interviews on Nigeria & Uganda but eSurvey suggests limited accessibility<br>**Key barriers**<br>• HIVST is reported to be available OTC at retail pharmacies in Nigeria, but it is unclear about utilization or linkage to post-test services. Uptake seems to remain low & data shows concerns over accuracy & availability of follow-up care.<br>**Key Drivers**<br>• HIVST has been in policy documents for several years already. It is one of the most advanced SC tools for STIs available. Recommended since 2017 in Kenya & Nigeria; addressed in the most recent SC national guidelines in Nigeria & Uganda. |
| **REC 24: SRHR empowerment for women living with HIV (WLWH)** | **Desktop research**<br>• Psychological stress among WLWH can compromise linkage & retention to HIV care, reduced health-related quality of life, increased risk of HIV transmission & depression (54). Self-efficacy & empowerment are important predictors of treatment outcomes & general wellbeing.<br>• The prevalence of HIV infections among women remains high in Kenya, Nigeria & Uganda.<br>• Local case studies & interventions in Kenya & Nigeria showed positive impact.<br>**Sentiment analysis**<br>• eSurvey reports viability in Kenya, moderate availability in Nigeria & Uganda,<br>**Key barriers**<br>• Lack of data & national programs to support this policy<br>**Key Drivers**<br>• Addressed in the 2012 Guidelines for Prevention of Mother to Child Transmission (PMTCT) of HIV/AIDS in Kenya & in Nigeria's 2020 National Guidelines on SC. Not addressed in Uganda's 2020 SC guidelines |

### REFRENCES


- 1. S.O.U.L. foundation. Ensuring every woman can deliver with dignity Uganda: S.O.U.L. foundation; 2020 [Available from: https://www.souluganda.org/maternalhealth#:~:text=Uganda's%20Maternal%20Mortality%20Rate%20(MMR,related%20to%20pregnancy%20or%20delivery.
- 2. Keats E, Akseer N, Bhatti Z, Macharia W, Ngugi AK, Rizvi A, et al. Assessment of Inequalities in Coverage of Essential Reproductive, Maternal, Newborn, Child, and Adolescent Health Interventions in Kenya. JAMA Network Open. 2018;1:e185152.
- 3. MOH. Guidelines on Maternal Nutrition in Uganda. Uganda: MINISTRY OF HEALTH; 2010. p. 37.
- 4. Sanitation MoPHa. National Guidelines for quality obstetrics and perinatal care 2010. In: Sanitation MoPHa, editor. Kenya: Ministry of Public Health and Sanitation; 2010. p. 410.
- 5. Hensley JG. Leg cramps and restless legs syndrome during pregnancy. Journal of midwifery & women's health. 2009;54(3):211-8.
- 6. Lee KA, Gay CL. Sleep in late pregnancy predicts length of labor and type of delivery. American journal of obstetrics and gynecology. 2004;191(6):2041-6.
- 7. Rondó PH, Ferreira RF, Nogueira F, Ribeiro MC, Lobert H, Artes R. Maternal psychological stress and distress as predictors of low birth weight, prematurity and intrauterine growth retardation. European journal of clinical nutrition. 2003;57(2):266-72.
- 8. Marcus SM, Flynn HA, Blow FC, Barry KL. Depressive symptoms among pregnant women screened in obstetrics settings. Journal of women's health. 2003;12(4):373-80.
- 9. Dayan J, Creveuil C, Herlicoviez M, Herbel C, Baranger E, Savoye C, et al. Role of anxiety and depression in the onset of spontaneous preterm labor. American journal of epidemiology. 2002;155(4):293-301.
- 10. Vermani E, Mittal R, Weeks A. Pelvic girdle pain and low back pain in pregnancy: a review. Pain Practice. 2010;10(1):60-71.
- 11. de Groot N, Birnie E, Vermolen JV, Dorscheidt JJ, Bonsel GJ. Replication Data for: The prevalence of adverse postnatal outcomes for mother and infant in the Netherlands. 2019.
- 12. Johnson P, Mount K, Graziano S. Functional bowel disorders in pregnancy: effect on quality of life, evaluation and management. Acta obstetricia et gynecologica Scandinavica. 2014;93(9):874-9.
- 13. Darvall K, Bate G, Adam D, Bradbury A. Generic health-related quality of life is significantly worse in varicose vein patients with lower limb symptoms independent of CEAP clinical grade. European Journal of Vascular and Endovascular Surgery. 2012;44(3):341-4.
- 14. Organization WH. WHO recommendations for augmentation of labour: World Health Organization; 2014.
- 15. Nendela A, Richens Y, Wakasiaka S, Omoni G. Fear of birth in Kenya and the UK: Differences and similarities. African Journal of Midwifery and Women's Health. 2018;12(4):201-4.
- 16. World Health Organization. Self-administration of injectable contraception 2020 [Available from: https://www.who.int/reproductivehealth/self-care-interventions/injectable-contraception/en/.

- 17. Corroon M, Kebede E, Spektor G, Speizer I. Key Role of Drug Shops and Pharmacies for Family Planning in Urban Nigeria and Kenya. Glob Health Sci Pract. 2016;4(4):594-609.
- 18. PATH. DMPA-SC evidence to practice meeting: Increasing access, empowering women meeting report: Nairobi, kenya. 2018:42.
- 19. Olawo AA, Bashir I, Solomon M, Stanback J, Ndugga BM, Malonza I. "A cup of tea with our CBD agent … ": community provision of injectable contraceptives in Kenya is safe and feasible. Glob Health Sci Pract. 2013;1(3):308-15.
- 20. Hu E, Ikeako L, Obiora-Okafor N. The use of depot medroxyprogesterone acetate (DMPA) injectable contraceptive in Enugu, Nigeria. Nigerian journal of medicine: journal of the National Association of Resident Doctors of Nigeria. 2012;21(3):266-71.
- 21. Cover J, Namagembe A, Tumusiime J, Lim J, Cox CM. Ugandan providers' views on the acceptability of contraceptive self-injection for adolescents: a qualitative study. Reprod Health. 2018;15(1):165.
- 22. Performance Monitoring For Action. Uganda, Round 6: Family Planning Brief. 2018.
- 23. Biddlecom AE, Munthali A, Singh S, Woog V. Adolescents' views of and preferences for sexual and reproductive health services in Burkina Faso, Ghana, Malawi and Uganda. African journal of reproductive health. 2007;11(3):99.
- 24. World Health Organization. Making health services adolescent friendly: Developing national quality standards for adolescent friendly health services. 2012.
- 25. Gay J. High-Impact Practices in Family Planning (HIPs),“. Adolescent-Friendly Contraceptive Services: Mainstreaming Adolescent-Friendly Elements Into Existing Contraceptive Services.
- 26. Kennedy CE, Yeh PT, Gonsalves L, Jafri H, Gaffield ME, Kiarie J, et al. Should oral contraceptive pills be available without a prescription? A systematic review of over-the-counter and pharmacy access availability. BMJ Global Health. 2019;4(3):e001402.
- 27. Plus S. Regulation of Drug Shops and Pharmacies Relevant to Family Planning: A Scan of 32 Developing Countries. Maryland, USA; 2017.
- 28. Riley P, Sean C, Dalious M. Regulation of Drug Shops and Pharmacies Relevant to Family Planning: A Scan of 32 Developing Countries. Sustaining Health Outcomes through the Private Sector Plus Project (SHOP Plus projects); 2017. p. 84.
- 29. World Health Organization. Home-based ovulation predictor kits 2020 [Available from: https://www.who.int/reproductivehealth/self-care-interventions/ovulation-predictor-kits/en/.
- 30. Yeh PT, Kennedy CE, Van der Poel S, Matsaseng T, Bernard L, Narasimhan M. Should home-based ovulation predictor kits be offered as an additional approach for fertility management for women and couples desiring pregnancy? A systematic review and meta-analysis. BMJ Global Health. 2019;4(2):e001403.
- 31. World Health Organization. HIV/AIDS 2020 [Available from: https://www.who.int/hiv/mediacentre/news/condoms-joint-positionpaper/en/.
- 32. Kosugi H, Shibanuma A, Kiriya J, Wafula SW, Jimba M. Consistent condom use among highly effective contraceptive users in an HIV-endemic area in rural Kenya. PLoS One. 2019;14(5):e0216208.
- 33. De Cock KM, Rutherford GW, Akhwale W. Kenya AIDS Indicator Survey 2012. J Acquir Immune Defic Syndr. 2014;66 Suppl 1:S1-2.

- 34. Dumo E. Despite recession, condoms sell very well – Lagos Kiosk owners: PUNCH; 2017 [Available from: https://punchng.com/despite-recession-condoms-sell-very-well-lagos-kiosk-owners/.
- 35. The New York Times. Nigeria Tested by Rapid Rise in Population 2012 [Available from: https://www.nytimes.com/2012/04/15/world/africa/in-nigeria-a-preview-of-an-overcrowded-planet.html.
- 36. Daily Monitor. Condom use on the rise - minister: Daily Monitor; 2018 [Available from: https://www.monitor.co.ug/uganda/news/national/condom-use-on-the-rise-minister-1740242.
- 37. De Coninck Z, Marrone G. Trends and determinants of condom use in Uganda. East Afr J Public Health. 2012;9(3):105-11.
- 38. World Health Organization. Condom and Lubricant Programming. 2013.
- 39. aidsmap. Consistent and correct condom use reduces risk of bacterial STIs by 60% 2012 [Available from: https://www.aidsmap.com/news/sep-2012/consistent-and-correct-condom-use-reduces-risk-bacterial-stis-60.
- 40. Okon A. 70% of lubricants in Nigeria are substandard — SON Nigeria: PUNCH; 2018 [Available from: https://punchng.com/70-of-lubricants-in-nigeria-are-substandard-son/.
- 41. Mehra D, Östergren P-O, Ekman B, Agardh A. Inconsistent condom use among Ugandan university students from a gender perspective: a cross-sectional study. Global Health Action. 2014;7(1):22942.
- 42. Stillman M, Owolabi O, Fatusi AO, Akinyemi AI, Berry AL, Erinfolami TP, et al. Women's self-reported experiences using misoprostol obtained from drug sellers: a prospective cohort study in Lagos State, Nigeria. BMJ Open. 2020;10(5):e034670.
- 43. Fabic MS, Choi Y, Bongaarts J, Darroch JE, Ross JA, Stover J, et al. Meeting demand for family planning within a generation: the post-2015 agenda. Lancet. 2015;385(9981):1928-31.
- 44. Kabiru CW, Ushie BA, Mutua MM, Izugbara CO. Previous induced abortion among young women seeking abortion-related care in Kenya: a cross-sectional analysis. BMC Pregnancy Childbirth. 2016;16:104.
- 45. Maina BW, Mutua MM, Sidze EM. Factors associated with repeat induced abortion in Kenya. BMC Public Health. 2015;15(1):1048.
- 46. Unemo M, Bradshaw CS, Hocking JS, de Vries HJC, Francis SC, Mabey D, et al. Sexually transmitted infections: challenges ahead. The Lancet Infectious Diseases. 2017;17(8):e235-e79.
- 47. Korenromp EL, Rowley J, Alonso M, Mello MB, Wijesooriya NS, Mahiané SG, et al. Global burden of maternal and congenital syphilis and associated adverse birth outcomes—Estimates for 2016 and progress since 2012. PLOS ONE. 2019;14(2):e0211720.
- 48. WHO. Sexually transmitted infections (STIs) 2019 [Available from: https://www.who.int/news-room/fact-sheets/detail/sexually-transmitted-infections-(stis).
- 49. WHO. HIV/AIDS 2020 [Available from: https://www.who.int/news-room/fact-sheets/detail/hiv-aids.
- 50. WHO. Human papillomavirus (HPV) and cervical cancer 2020 [Available from: https://www.who.int/news-room/fact-sheets/detail/human-papillomavirus-(hpv)-and-cervical-cancer.
- 51. Campos NG, Tsu V, Jeronimo J, Njama-Meya D, Mvundura M, Kim JJ. Cost-effectiveness of an HPV self-collection campaign in Uganda: comparing models for delivery

of cervical cancer screening in a low-income setting. Health Policy and Planning. 2017;32(7):956-68.
- 52. Rowley J, Vander Hoorn S, Korenromp E, Low N, Unemo M, Abu-Raddad LJ, et al. Chlamydia, gonorrhoea, trichomoniasis and syphilis: global prevalence and incidence estimates, 2016. Bull World Health Organ. 2019;97(8):548-62P.
- 53. Organization WH. WHO consolidated guideline on self-care interventions for health: sexual and reproductive health and rights. 2019:153.
- 54. Adamu A, McHunu G, Naidoo J. Stress and resilience among women living with HIV in Nigeria. African Journal of Primary Health Care & Family Medicine. 2019;11.
- 55. MoH. Kenya National Cancer Screening Guidelines. 2018.
- 56. MoH. HIV SELF-TESTING. An operational manual for the delivery of HIV Self-Testing services in Kenya. 2017.
- 57. MoH. Guidelines for Prevention of Mother to Child Transmission (PMTCT) of HIV/AIDS in Kenya. 2012.
- 58. MoH. National Family Planning Guidelines for Service Providers. In: Reproductive and Maternal Health Services Unit (RMHSU) DoFH, editor. 6th ed2018.
- 59. MoH. National Guidelines for Quality Obstetrics and Perinatal Care
- 60. Services MoM. STANDARDS and GUIDELINES for reducing morbidity & mortality from unsafe abortion in Kenya. 2012.
- 61. Ministry of Public Health and Sanitation. National Road Map for accelerating the attainment of the mdgs related to maternal health and newborn health in kenya. In: Sanitation MoPHa, editor. Kenya: Ministry of Public Health and Sanitation; 2010.
- 62. Sanitation MoPHa. NATIONAL GUIDELINES ON ESSENTIAL NEWBORN CARE. In: Sanitation MoPHa, editor. Kenya: Ministry of Public Health and Sanitation; 2015. p. 70.
- 63. Ministry of Health (FMoH). ACCELERATED REDUCTION OF MATERNAL AND NEONATAL MORTALITY IN NIGERIA: The Road Map for Action 2019-2021. Nigeria: Ministry of Health; 2019-2021. p. 80.
- 64. Federal Ministry of Health. Antenatal care: An Orientation Package for Health Care Providers. Nigeria: Ministry of Health; 2017. p. 101.
- 65. FEDERAL MINISTRY OF HEALTH. NATIONAL GUIDELINE FOR THE INTRODUCTION AND SCALE-UP OF DMPA-SC SELF-INJECTION. 2019.
- 66. FEDERAL MINISTRY OF HEALTH. Family Planning Services – National Family Planning Communication Plan, Strategy for Increasing the use of Modern Contraceptives in Nigeria 2020.
- 67. FMOH. HIV Self-Testing. Operational Guidelines for the Delivery of HIV Self-Testing in Nigeria. 2018.
- 68. Ajenifuja OK, Ikeri NZ, Adeteye OV, Banjo AA. Comparison between self sampling and provider collected samples for Human Papillomavirus (HPV) Deoxyribonucleic acid (DNA) testing in a Nigerian facility. The Pan African Medical Journal. 2018;30.
- 69. FMOH. Family Planning On-The-Job Training Manual, Facilitator’s Manual Federal Ministry of Health, Nigeria; 2019.
- 70. FMOH. Manual for training doctors and nurse/midwives on long-acting reversible contraceptive (LARC) methods (IUDs and Contraceptive Implants). 2015.
- 71. Federal Ministry of Health Nigeria. National Health Promotion Policy. 2019.
- 72. The National Agency for the Prohibition of Trafficking in Persons (NAPTIP). Violence Against Persons (Prohibition) Act 2018.

- 73. Federal Ministry of Health Nigeria. National strategic frame work for the elimination of obstetric fistula in nigeria 2019 - 2023. In: Health Mo, editor. Nigeria: Ministry of Health. p. 65.
- 74. Ferguson L, Fried S, Matsaseng T, Ravindran S, Gruskin S. Human rights and legal dimensions of self care interventions for sexual and reproductive health. bmj. 2019;365.
- 75. Division P. Essential Medicines and Health Supplies List for Uganda. Ministry of Health; 2016.
- 76. MoH. National Policy Guidelines for Sexual and Reproductive Health services. Reproductive Health division, Department of Community Health, MOH; 2012.
- 77. PATH TBM. Lesson 10: How to counsel clients on DMPA-SC self-injection.
- 78. Nakisige C, Trawin J, Mitchell-Foster S, Payne BA, Rawat A, Mithani N, et al. Integrated cervical cancer screening in Mayuge District Uganda (ASPIRE Mayuge): a pragmatic sequential cluster randomized trial protocol. BMC public health. 2020;20(1):142.
- 79. RHITES–EC U. Engaging a Key Population Drop-In Center to Increase Uptake of HIV Self-Testing Services: The Experience of Busia Health Center IV in East Central Uganda2020.
- 80. Esther Nasuuna FN, James Wanyama, Alice Namale, Martin Ssuuna, Alex Muganzi, Joanita Kigozi. HIV Self testing among Key populations and sexual partners of new mtohers in Uganda. 2020.
- 81. USAID. HIV Self-Test Targeted Distribution and Linkage Plan Summary. 2020.
- 82. Facing Uganda’s law on abortion - Experiences from Women & Service Providers. Uganda; 2016.
- 83. Uganda Condom Market Champion.
- 84. Health TRoUMo. Uganda Clinical Guidelines 2016 – National Guidelines for Management of Common Conditions. 2016.
- 85. The Republic of Uganda Ministry of Health Acp. ADDENDUM TO THE HIV TESTING SERVICES POLICY & IMPLEMENTATION GUIDELINES HIV SELF-TESTING & ASSISTED PARTNER NOTIFICATION SERVICES. Uganda; 2018.
- 86. Theory of Self-injection Scale-up.
- 87. Health UMo. Strategic Plan for Cervical Cancer Prevention and Control in Uganda, 2010–2014. Uganda; 2010.
- 88. Ministry of Health. Uganda National Self-care guidelines for Sexual and Reproductive health and Rights. In: Health Mo, editor. Uganda: Ministry of Health; 2020. p. 52.
- 89. MOH. HIV self-testing & HIV assisted partner notification services policy & implementation guidelines 2018 : Addendum to 2016 HTS policy & implementation guidelines. Uganda: MINISTRY OF HEALTH; 2018. p. 67.
- 90. MOH. Adolescent Health Policy Guidelines andService Standards. In: Reproductive Health division DoCH, MOH, editor. Kampala: Ministry of health; 2012.
- 91. AU. How to take your own sample for a HPV test: Self-collection instructions to be provided only by a healthcare provider during a consultation. In: Health Do, editor. Australia: Australia Government, Department of Health; 2017.
- 92. Australia Government. Quick Reference Guide - Self-Collected Vaginal Sample for HPV Test. In: Health Do, editor. Australia: Australia Government; 2020.
- 93. Uganda. Constitution of the Republic of Uganda: Uganda Print. and Publishing Corporation; 1995.

- 94. PATH. DMPA-SC Self-injection virtual TOT. Uganda; 2020.
- 95. Ministry of Health of Uganda. Module 7 HPV Detection Tests & Sample Collection Techniques for HPV Testing. 2019.
- 96. PATH. DMPA-SC Self-injection virtual training for partners. 2020.
- 97. THE PENAL ACT. CHAPTER 120, (1950).
- 98. Ministry of Health of Uganda. Provision of DMPA-SC Self-Injection Circular. 2020.
- 99. PATH. Proposed Self-injection guidelines for Uganda 2020.
- 100. Observation checklist to assess health workers orienting clients on DMPA-SC self-injection. 2021.
- 101. Ministry of Health of Uganda. The National Policy Guidelines and Service Standards for Sexual and Reproductive Health and Rights. 2012.
- 102. Nankumbi J, Ngabirano TD, Nalwadda G. Maternal Nutrition Education Provided by Midwives: A Qualitative Study in an Antenatal Clinic, Uganda. Journal of Nutrition and Metabolism. 2018;2018:3987396.
- 103. Asiimwe G. C-section deliveries in Uganda; Why you should get concerned. Uganda: Makerere University's Institutional Repository; 2020.
- 104. Jebet J, Omoni G, Waithira M. Health Education on Birth Preparedness: What is Happening? a Study Among Mothers Attending Antenatal Clinics in Migori Country, Kenya. International Journal of Nursing Care. 2017;5:5.
- 105. Moh U. Guidelines on Maternal Nutrition in Uganda. Ministry of Health, Uganda. 2010.
- 106. Bryce J, Coitinho D, Darnton-Hill I, Pelletier D, Pinstrup-Andersen P, Maternal, et al. Maternal and child undernutrition: effective action at national level. The Lancet. 2008;371(9611):510-26.
